# Exome sequencing and analysis of 44,028 British South Asians enriched for high autozygosity

**DOI:** 10.1101/2025.06.05.25329068

**Authors:** Hye In Kim, Christopher DeBoever, Klaudia Walter, Georgios Kalantzis, Chen Li, Sahar V. Mozaffari, Kousik Kundu, Benjamin M. Jacobs, Pedrum Mohammadi-Shemirani, Anthony M. Musolf, Jonathan M. Davitte, Melis A. Aksit, Joseph Gafton, Katrina Catalano, Adem Y. Dawed, Robert R. Graham, Bin Guo, Namrata Gupta, Teng Hiang Heng, Karen A. Hunt, Vivek Iyer, Claudia Langenberg, Frederik H. Lassen, Daniel G. MacArthur, Eamonn R. Maher, Cyrielle Maroteau, William Newman, Stephen O’Rahilly, Duncan S. Palmer, Iaroslav Popov, Moneeza Siddiqui, Michael Simpson, Marie Spreckley, John Wright, Genes and Health Research Team, Guillermo del Angel, Slavé Petrovski, Emily R. Holzinger, Joseph C. Maranville, Laura Addis, Richard M. Turner, Karol Estrada, Simone Longerich, Joanna M. M. Howson, Yalda Jamshidi, Eric B. Fauman, Melissa R. Miller, Dorothée Diogo, Richard C. Trembath, Sarah Finer, Hilary C. Martin, David A. van Heel

## Abstract

Genes and Health (G&H) is a biomedical study of adult British-Pakistani and -Bangladeshi research volunteers enriched for autozygosity. We performed whole exome sequencing in 44,028 G&H participants, establishing the largest publicly available South Asian exome resource linked to longitudinal electronic health records. We performed association analyses for 646 traits under additive and recessive models, and meta-analysis of 33 cardiometabolic traits with UK Biobank, finding more than 100 novel gene-phenotype associations such as *ADAM15* with pulmonary oedema and *ADCY6* with intracerebral haemorrhage. We identified 2,991 genes with rare biallelic predicted loss-of-function (“knockout”) genotypes, 546 of which had not been previously reported. We show that the presence of knockouts in adults is associated with 2.2-times higher likelihood of drugs progressing beyond Phase 1 clinical trial. We further illustrate how their phenotypic profile can enhance efficacy and safety assessment of drug targets and aid in the interpretation of variants with ambiguous clinical significance in autosomal recessive disease genes.

## Introduction

Major advances in our understanding of human diseases have been achieved through genotyping, sequencing, and analyses of large population-or hospital-based cohorts^1–5^. However, there remains a critical underrepresentation of non-European ancestral groups in genetic datasets^6,7^. Addressing this imbalance is essential not only for ensuring equity in genetic research but also for maximizing discoveries harnessing distinct variant spectra present in diverse ancestry groups^8–11^.

Genes & Health (G&H) is a population-based cohort study of British-Bangladeshi and -Pakistani adults, aiming to improve the health outcomes of these communities through genetic research. G&H benefits from comprehensive lifelong health care data from the UK National Health Service, which allows the systematic extraction of disease diagnoses and clinical traits for genetic association analyses^12,13^ and the detailed medical record review in specific carriers of interest^14,15^.

A distinctive characteristic of this cohort is its high degree of autozygosity, and hence enrichment of homozygous genotypes of rare variants^16,17^. Particularly informative are homozygous carriers of loss-of-function variants, often called “human knockouts”. Studies from this cohort and others have illustrated the valuable insights human knockouts can provide into human biology, disease mechanisms, and drug development^14,16,18–20^.

In this study, we present the analyses of whole exome sequences of 44,028 participants in G&H, representing the largest South Asian exome resource to date with linked electronic medical data. We share key findings from rare variant association analyses, using both additive and recessive models, alongside a meta-analysis with UK Biobank that uncovers numerous novel gene-phenotype associations. We further characterize genes with human knockouts in this cohort and highlight different insights they provide from biological, drug development, and clinical perspectives.

## Results

### Protein coding variation and population structure in 44,028 South Asian exome sequences

We identified a total of 4,723,926 variants (4,458,984 SNVs and 264,942 INDELs) from 44,028 exome sequences after stringent quality control (**Supplementary Methods**). Across 17,545 MANE transcripts^21^, we found 122,690 predicted loss-of-function (pLoF) variants that are high-confidence (pLoF-HC) by LOFTEE^22^, and 1,704,012 missense variants, of which 524,290 are predicted damaging missense (pDM) (CADD>20, Polyphen2>0.445, and SIFT deleterious) (**Figure 1A**). As expected, pLoF and pDM variants were heavily enriched amongst singleton and ultra-rare variants (**Figure 1B**). We compared the allele frequency of the variants in G&H to those in the Genome Aggregation Database^22^ (gnomAD, v4.1) which catalogues variants from 807,162 genomes and exomes of diverse ancestry, including 45,546 of South Asian ancestry. Of all the variants in G&H, 26.2% are absent from gnomAD, a further 24.4% are in gnomAD but not in the non-Finnish European (NFE) subset, and a further 18.8% have >10-fold higher allele frequency in G&H compared to gnomAD NFE (**Figure 1C**). 325,276 variants were significantly enriched in a subset of 17,172 unrelated individuals from G&H (**Methods**) compared to gnomAD-NFE (Fisher’s exact test p<4.93×10^−8^ with Bonferroni correction).

**Figure 1.**
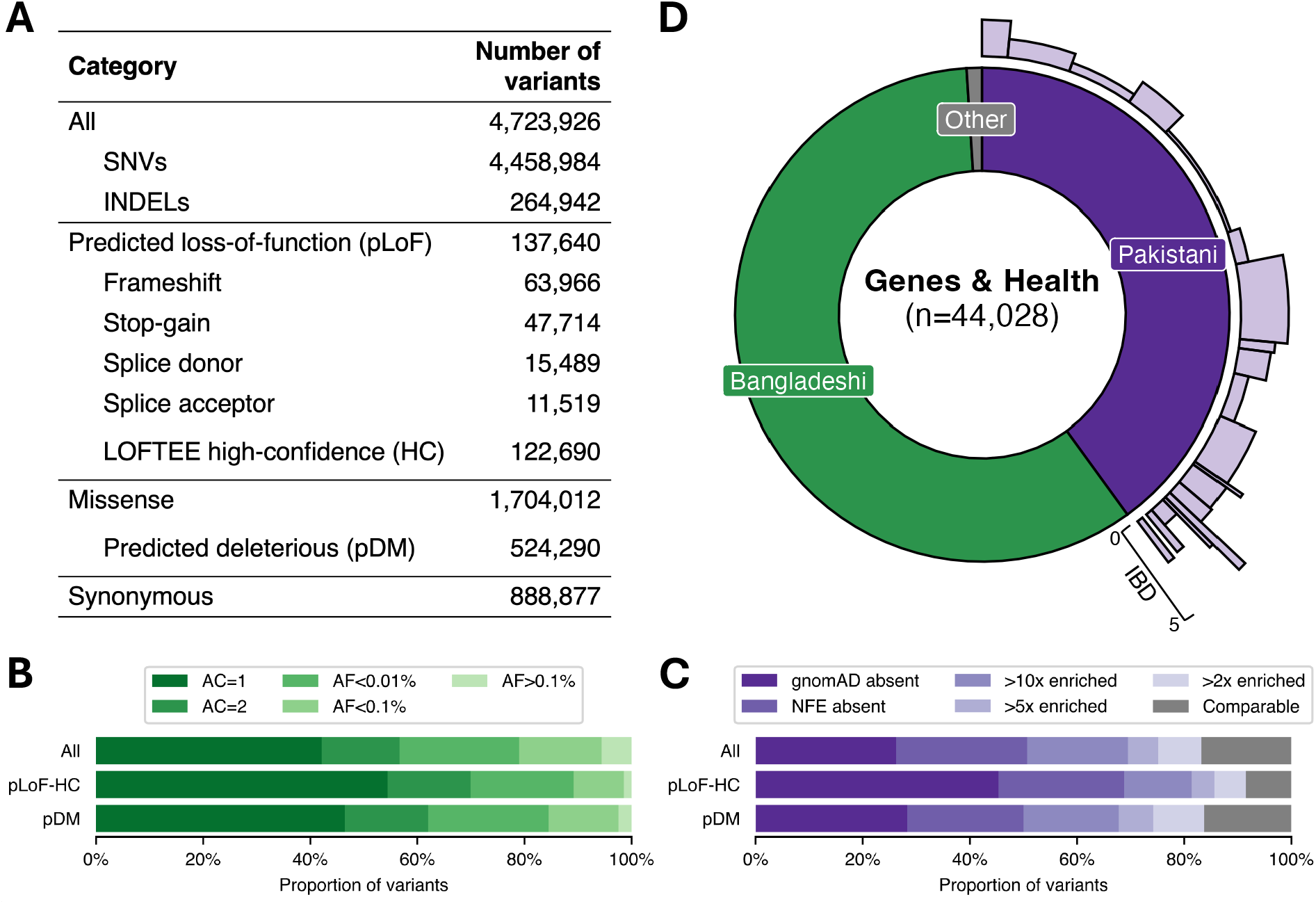
Protein coding variation and population structure in 44,028 Genes & Health exomes. (A) Number of all variants, SNVs and INDELs, and per functional category. pLoF-HC = high confidence predicted loss-of-function; pDM = predicted deleterious missense. (B) Allele frequency (AF) spectrum of all, pLoF-HC, and pDM variants. (C) Proportions of variants completely absent in gnomAD; absent in the non-Finnish Europeans (NFE) subset of gnomAD; with 10x, 5x or 2x greater allele frequency compared to gnomAD NFE. The grey portion includes variants in both G&H and gnomAD with comparable allele frequency. (D) South Asian ancestry breakdown in G&H and sub-population structure among British-Pakistanis. For the British-Pakistani sub-populations in light purple, the width indicates the relative proportion of clusters, and the height indicates the IBD score in the clusters.

G&H consists of British residents of self-identified Pakistani (40%) and Bangladeshi (59%) ancestry (**Figure 1D**). Principal components analysis (PCA) demonstrates that there is considerable population structure among the British-Pakistanis but much less among the British-Bangladeshis^23^ (**Supplementary Figure 1**). The population structure of British-Pakistanis is strongly influenced by the biraderi social stratification system and is characterized by extensive identity-by-descent (IBD) sharing due to founder effects^24^. Using an IBD-based clustering method^24^ (**Methods**), we identified 21 clusters among 8,109 unrelated British Pakistani individuals (**Figure1D; Supplementary Figures 1 and 2; Supplementary Table 1**) representing putative subgroups. Several clusters have particularly extensive IBD sharing (**Supplementary Figure 2**). We identified 15,202 variants that were significantly enriched in specific clusters compared to all the others combined, which may have resulted from founder events or possibly positive selection, described further in **Supplementary Note 1 (Supplementary Figures 2 and 3; Supplementary Tables 2 and 3**).

**Figure 2.**
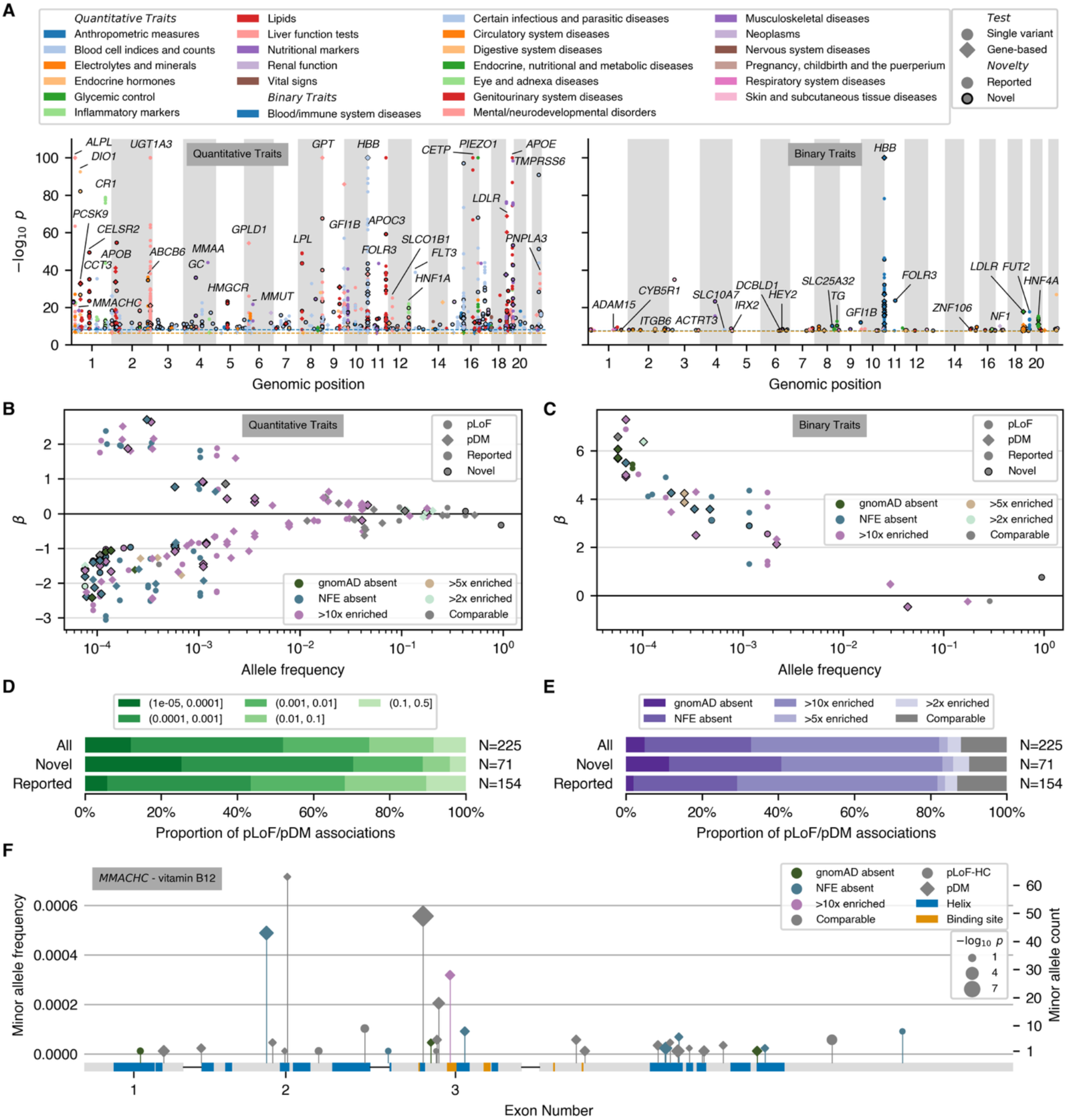
Rare variant association analyses. (A) Manhattan plots showing significant quantitative trait (left) and binary trait (right) associations by non-olfactory and non-MHC variants/genes. Synonymous gene-based associations and associations that were not significant after conditioning on nearby GWAS associations are omitted. Fill colour: phenotype categories; circle markers: single variant associations; diamond markers: gene-based associations. Dark marker outlines indicate that the gene-phenotype association is annotated as novel. −log10 p-values were truncated at 100. Blue dashed lines: single variant p-value cutoffs; orange dashed lines: gene-based p-value cutoffs. (B-C) Single variant MAF versus effect size (*β*) for significant QT (B) and BT (C) associations. Circle markers: pLoF variants; diamond markers: pDM variants; Dark marker outlines: novel gene-phenotype association. Purple marker colours: G&H MAF enrichment. (D) Proportion of pLoF and pDM single variant associations annotated as novel or previously reported, stratified by MAF bin. (E) Proportion of pLoF and pDM single variant associations annotated as novel or previously reported, stratified by allele frequency ratio between G&H and gnomAD NFE. (F) pLoF-HC and pDM variants in *MMACHC* with MAF < 0.001 that were included in the gene-based test with significant association with vitamin B12 levels. Purple marker colours: G&H MAF enrichment; circle markers: pLoF variants; diamond markers: pDM variants. Blue and orange boxes indicate Uniprot alpha helices and binding sites, respectively. Marker size indicates −log10 association p-value of each variant.

**Figure 3.**
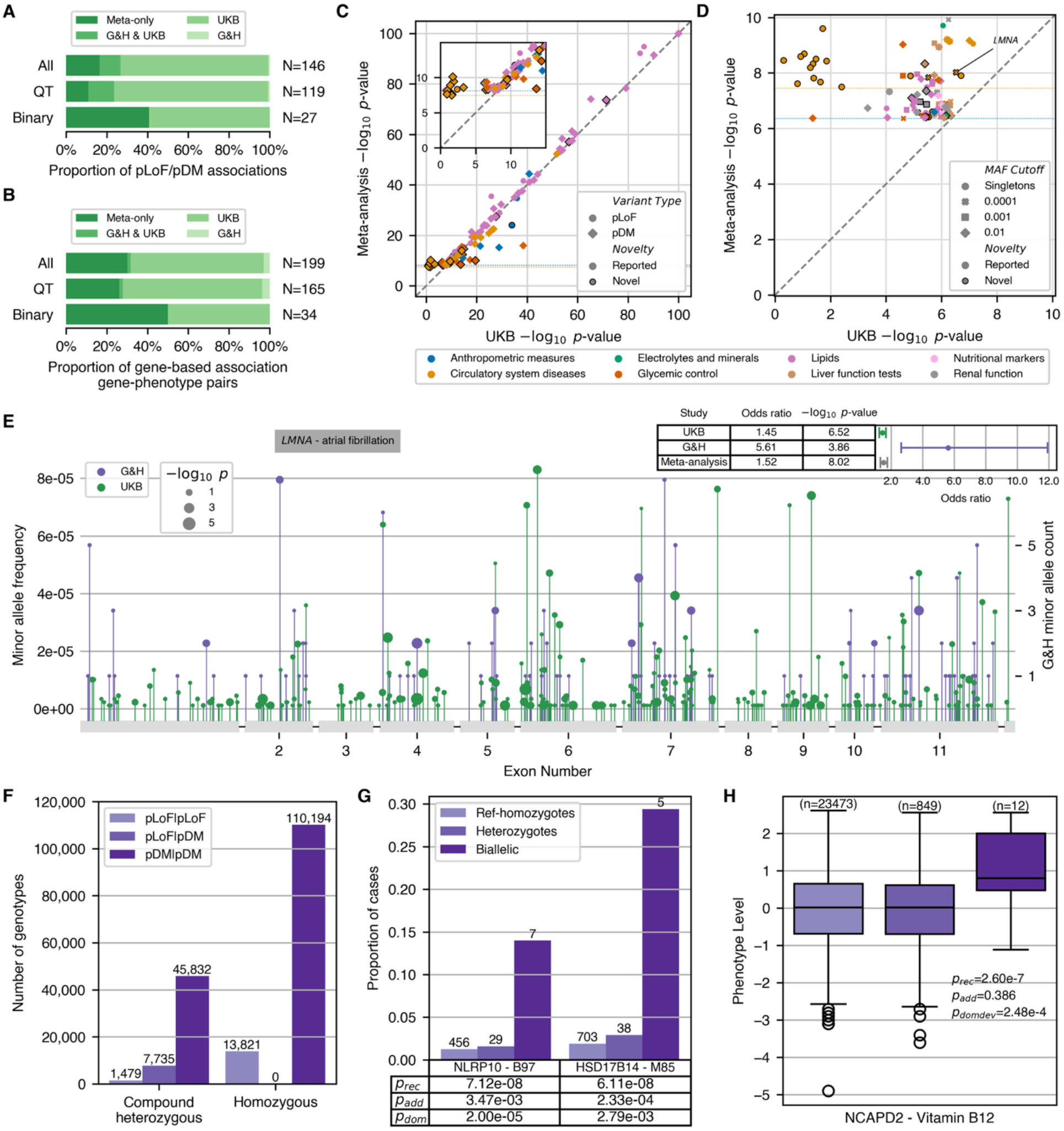
Meta-analysis and recessive burden analyses. (A) Proportion of pLoF and pDM single variant associations and (B) proportion of gene-phenotype pairs with gene-based associations stratified by the significance status in respective studies: only in the meta-analysis; in both G&H and UKB; in UKB only; or in G&H only. (C) Meta-analysis −log10 p-value versus UKB −log10 p-value for the significant single variant pLoF/pDM associations from meta-analysis. −log10 p-values were truncated at 100. Marker colour indicates phenotype categories. Marker shape indicates variant type. Dark marker outline indicates that the gene-phenotype association is annotated as novel. Blue and orange dashed lines indicate QT and BT p-value cutoffs, respectively. (D) Meta-analysis −log10 p-value versus UKB −log10 p-value for the significant non-MHC, non-olfactory, and nonsynonymous gene-based associations from meta-analysis. Marker shape indicates variant frequency cutoff used for the burden. (E) pLoF and missense variants with MAF<0.0001 in the *LMNA* burden associated with atrial fibrillation. Purple and green marker colours indicate variants in G&H or UKB, respectively. Marker size indicates −log10 association p-value of each variant. (F) Forest plot showing the odds ratios and p-values from G&H, UKB, and meta-analysis for the gene-based association between *LMNA* and atrial fibrillation. (F) Counts of compound heterozygous and homozygous genotypes found among the 39,148 phased exomes stratified by variant consequence. (G) Bar plots showing the proportion of diseased individuals per burden genotype of pLoF and pDM variants for significant recessive associations with binary traits. The number of diseased carriers is shown at the top of each bar and p-values for recessive, additive, and dominance deviation tests are shown in the inset table. (H) Box plots showing the quantile normalized levels of covariate adjusted phenotypes per burden genotype of pLoF and pDM variants for a significant recessive association with a quantitative trait. Number of carriers are indicated at the top of each box.

There were 8,450 variants across 2,855 genes that were curated as pathogenic or likely pathogenic (PLP) in ClinVar^25^ and had at least one heterozygous or homozygous genotype in G&H exomes (**Supplementary Note 2; Supplementary Figure 4A; Supplementary Table 4**). Among the 81 clinically actionable genes defined by the American College of Medical Genetics and Genomics^26^ (ACMG SF v3.2), we found 1,012 individuals heterozygous for PLP variants in autosomal dominant genes and 7 individuals homozygous for PLP variants in autosomal recessive genes. (**Supplementary Note 2; Supplementary Table 4)**. Compared to a size-matched subset of European-ancestry exomes from UK Biobank (UKB-EUR), a smaller proportion of pLoF and pDM variants in G&H were already present in ClinVar. However, a higher proportion of the ClinVar variants in UKB-EUR were annotated to be pathogenic or to have unknown significance or conflicting interpretations compared to those in G&H (**Supplementary Note 2; Supplementary Table 5**). These presumably reflect the relative paucity of patients of South Asian ancestry who have undergone clinical sequencing, as well as potential geographical differences in the practice of reporting variants to ClinVar.

**Figure 4.**
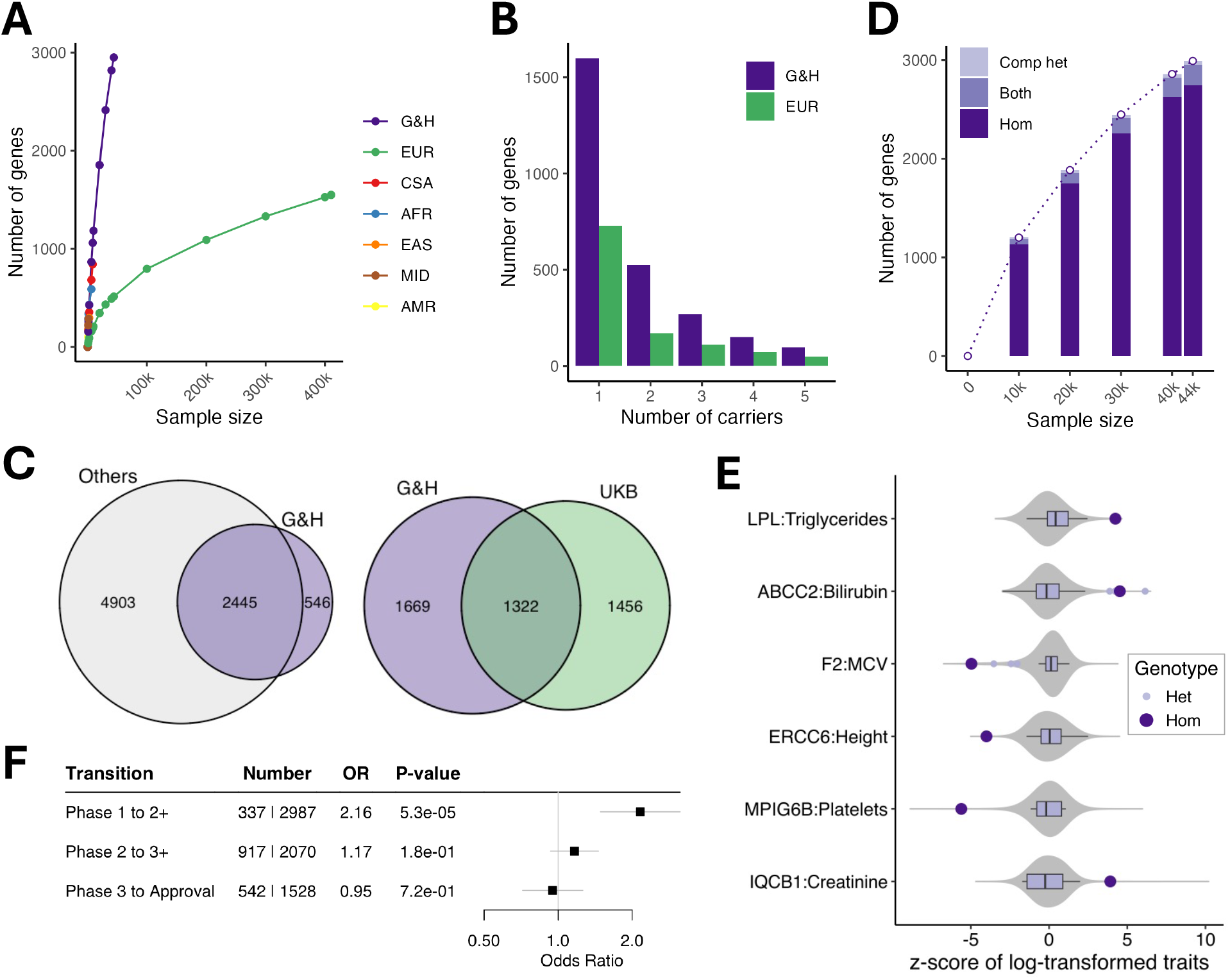
Identification of genes with human knockouts and insights into drug development and clinical variant interpretation. (A) Accrual of genes with at least one homozygous pLoF genotype in G&H and UK Biobank exomes stratified by ancestry groups. EUR, European; CSA, Central/South Asian; AFR, African; EAS, East Asian; MID, Middle Eastern; AMR, Admixed American ancestry. (B) Number of genes with up to 5 pLoF homozygous genotypes in 44,028 G&H exomes and 469,814 European-ancestry exomes from UK Biobank. (C) Overlap of genes with human knockouts in G&H exomes compared to those in four other genomic datasets (left) or to those in UK Biobank exomes (right). (D) Accrual of genes with at least one human knockout (either homozygous or compound heterozygous genotype) at increasing sample sizes of G&H exomes. (E) Disease-relevant quantitative phenotypes in the homozygous carriers of pLoF variants in AR disease genes. Large dots in dark purple indicate the lifetime median values of the homozygous carriers. Box plots in light purple show the distribution of values among heterozygous carriers. Grey violin plots show the distribution of the values among noncarriers. “Het” and “Hom” indicate heterozygous and homozygous carriers, respectively. (F) Enrichment analysis results of antagonistic drugs with human knockouts per trial phase transition.

### Rare variant association analyses

#### Exome-wide association analyses across 646 EHR-derived traits

We performed exome-wide association analyses using REGENIE^27^ for 55 quantitative traits (QTs) extracted from routine clinical and laboratory measurements and 591 binary traits (BTs) derived from diagnosis and clinical procedure codes (**Supplementary Table 6; Supplementary Methods**). To define significance thresholds, we permuted the phenotypes once at the second step of REGENIE and determined the p-value threshold corresponding to a false discovery rate (FDR) of 5% (**Supplementary Methods**). This approach ensures we control for the fine-scale population structure and relatedness in the sample. We observed minimal genomic inflation in the summary statistics, indicating that REGENIE was adequately controlling for these potential confounders (**Supplementary Note 3; Supplementary Table 7**). Overall, 267 of the 3,096 significant single-variant associations involved variants that were significantly enriched in specific British Pakistani sub-populations, and 82% of these associations were for HLA variants (**Supplementary Note 1**).

For QTs, we identified 1,139 significant (p<7.5×10^−9^) single variant associations involving 46 phenotypes and 860 variants, and 685 significant (p<4.4×10^−7^) gene-based associations involving 38 phenotypes and 85 genes after removing HLA variants/genes, synonymous gene masks, and associations that were not significant after conditioning on nearby GWAS associations (**Figure 2A left; Supplementary Tables 8 and 9**). We replicated many known associations including genes implicated in lipid metabolism and liver enzymes (**Supplementary Note 3**). In several cases, G&H expanded the allelic series for genes with known associations. For instance, we found 4 pLoF and pDM variants in *APOB* that are absent or extremely rare in gnomAD NFE and have significant associations with lipid traits in G&H that are unreported elsewhere (**Supplementary Note 3**).

For BTs, we identified 159 significant (p<3.3×10^−8^) single variant associations involving 75 phenotypes and 130 variants, and 68 significant (p<3.5×10^−8^) gene-based associations involving 18 phenotypes and 13 genes (**Figure 2A right; Supplementary Tables 8 and 9**), again replicating many known gene-disease associations (**Supplementary Note 3**). The *HBB* gene on chromosome 11 was a notable outlier in terms of the number of significant associations (**Figure 2A right**), accounting for 8 of the 22 unique gene-BT pairs implicated by gene-based tests and 4 of 25 pairs from pLoF/pDM associations. Mutations in this gene are a well-known cause of sickle cell disease^28^ and beta thalassemia^29^. Accordingly, our associated phenotypes included these as well as other anaemias and various blood, iron, bilirubin and lipid traits, consistent with previous reports^30–37^. These associations are driven by several *HBB* variants that are either private or enriched in G&H and in South Asian ancestry in general (**Supplementary Figure 5**), including two that were highly enriched in specific Pakistani sub-populations (**Supplementary Note 1**). Consistent with this, a comparison with a similarly sized study of European ancestry exomes from UK Biobank^38^ revealed that the abundance of *HBB*-mediated associations is only identified in the South Asian G&H cohort (**Supplementary Figure 5**).

We assessed whether gene-phenotype pairs with significant associations in G&H have previously been implicated by genome-wide association studies (GWAS) or rare variant association studies (RVAS) (**Methods**). The single variant and gene-based associations implicated 40 unique gene-phenotype pairs for QTs and 202 for BTs (**Supplementary Figure 6)**. Of these 242 gene-phenotype pairs, 95 (39%) did not have prior genetic associations, and we term these “novel” (**Supplementary Table 10**). Variants that are absent in gnomAD or enriched in G&H tended to be rarer and have larger effects on the phenotypes, driving many novel gene-phenotype relationships (**Figure 2B**,**C**). Specifically, among pLoF/pDM single variant associations, variants in lower minor allele frequency (MAF) bins were more likely to implicate novel gene-phenotype pairs (Cochran-Armitage test, z=4,47, p=7.83×10^−6^, **Figure 2D**), as were variants absent from gnomAD (Fisher’s exact test, OR=6.39, p=4.99×10^−3^, **Figure 2E**). We highlight two examples of novel associations that are driven by variants private to or enriched in G&H (more in **Supplementary Note 3**):

- Singleton pLoF-HC burden (p=1.18×10^−8^, OR=275.9) and pLoF-HC/pDM burden (p=1.07×10^−8^, OR=83.9) in *ADAM15* were associated with pulmonary oedema risk. *ADAM15* regulates endothelial permeability, integrin signaling, inflammation, and extracellular matrix remodeling — processes linked to this condition^39–41^. GWAS has also implicated *ADAM15* in varicose veins^42^, another disorder related to vascular remodeling and dysfunction^43^. We found 12 singleton pLoF-HC variants (8 absent from gnomAD) and 20 pDM variants (13 absent from gnomAD or NFE). Pulmonary oedema was present in 15% of *ADAM15* variant carriers versus 0.29% of non-carriers. The high odds ratios suggest these variants are effectively Mendelian but with moderate penetrance. Aside from a rare *ADAM15* missense variant (p.Arg288Cys) linked to macular degeneration in a small study^44^, no prior rare variant associations have been reported for *ADAM15*.
- Burden of pLoF and pDM variants in *MMACHC* was associated with vitamin B12 levels (p=1.61×10^−11^, *β* =0.49). *MMACHC* encodes a vitamin B12 transporter, and rare *MMACHC* mutations cause the vitamin B12 disorder methylmalonic aciduria and homocystinuria, cblC type^45^. While a noncoding variant 12kb upstream has been linked to homocysteine levels^46^, this is the first association of coding variants with vitamin B12 levels. Of the 33 variants in the burden, three are absent from gnomAD, eight from gnomAD NFE, and one is 187-fold more frequent in G&H compared to gnomAD NFE (allele frequency of 3.18×10^−4^ versus 1.70×10^−6^) (**Figure 2F**).

#### Meta-analysis of 33 cardiometabolic traits with UK Biobank

Cross-ancestry meta-analysis may boost statistical power for discovery by increasing the allelic diversity within a gene and the number of carriers. Because British-Bangladeshi and British-Pakistani communities have the highest prevalence of cardiometabolic disease in the UK^47^, we performed a meta-analysis for cardiometabolic traits between 44,028 G&H participants and 409,499 European-ancestry UK Biobank (UKB) participants (**Methods**).

From single variant analyses, we identified 1,738 significant associations with consistent direction between G&H and UKB (p<3.3×10^−8^ for BTs, p<7.5×10^−9^ for QTs), 99 of which were not identified in either dataset alone (**Supplementary Table 11**). There were 146 associations for pLoF or pDM variants, 24 of which only became significant in the meta-analysis (**Figure 3A**). From gene-based analyses, we identified 710 significant associations (p<3.5×10^−8^ for BTs, p<4.5×10^−7^ for QTs), comprising 199 unique gene-phenotype pairs **(Supplementary Table 12)**. Of these, 60 (17 BTs, 43 QTs) rose to significance only in the meta-analysis (**Figure 3B**). In both single variant and gene-based analyses, the benefit of meta-analysis was particularly prominent for binary traits, with 41% and 50% of the significant associations, respectively, being only identified in the meta-analysis.

To explore the degree of power gain G&H contributed to the meta-analysis, we compared the p-values from the meta-analysis to those from UKB alone (**Figure 3C,D**). The greatest power gain was seen for binary traits related to circulatory system diseases in both single variant (**Figure 3C**) and gene-based (**Figure 3D**) results. In fact, the associations for these circulatory system diseases had stronger p-values in G&H compared to UKB, despite the nearly 10-fold difference in sample size between the two cohorts (**Supplementary Figure 7**) and lower case prevalences in G&H compared to UKB (**Supplementary Table 6**). Both the frequency and effect size of the variants were greater in G&H compared to UKB (**Supplementary Figure 7**), likely contributing to the stronger p-values. This suggests that power gain in cross-ancestry meta-analysis can be influenced not only by the difference in variant frequency spectrum but also by the difference in variant penetrance between ancestry groups.

Of the 60 and 24 gene-phenotype pairs newly implicated by gene-based and single variant meta-analysis, 26 and 13 were novel, respectively. To highlight some examples:

- Burden of pLoF and pDM variants in *LMNA* is associated with atrial fibrillation and flutter (OR=1.52, p=9.5×10^−9^) driven by rich allelic diversity in G&H and UKB (**Figure 3E**). Interestingly, the effect size of the burden was much greater in G&H than in UKB (heterogeneity p=5.51×10^−4^) (**Figure 3F**). Smaller candidate gene studies have suggested a potential link between *LMNA* and lone atrial fibrillation^48^, but this is the first report from unbiased genetic association analyses.
- *ADCY6* singleton pLoF-HC burden increases risk of intracerebral haemorrhage (OR=326, p=3.7×10^−9^). *ADCY6* encodes a member of the adenylyl cyclase protein family and plays an important role in maintaining a homeostatic contractile state of smooth muscle cells in the vessel wall and in regulating blood pressure^49^, which may explain its association with intracerebral haemorrhage. Biallelic mutations (mostly missense and one splice donor) in this gene have been reported to cause a lethal congenital contractures syndrome^50^ (OMIM 616287).
- A rare stop-gain variant (chr14-22773945-G-A, p.Arg473Ter) in *SLC7A7* was associated with atherosclerosis (OR=74.28, p=4.7×10^−9^). This variant is pathogenic in ClinVar for lysinuric protein intolerance, a rare autosomal recessive genetic disorder caused by impaired metabolism of lysine. Dysregulation in lysine metabolism has been linked to cardiometabolic pathophysiology^51,52^, which may influence the risk of atherosclerosis.

#### Recessive burden analyses with biallelic genotypes

High autozygosity in G&H can provide greater statistical power for recessive association analyses which have been relatively less explored^53–56^. There were 13,821 and 110,194 homozygous genotypes for pLoF and pDM variants with MAF<5%, respectively (**Supplementary Table 13**). We performed statistical phasing (**Supplementary Methods; Supplementary Figure 8A**) to identify compound heterozygous genotypes^56^, further increasing the number of biallelic pLOF and pDM genotypes by 45% (**Figure 3F; Supplementary Table 13**). Individual genotypes were then collapsed into three gene burdens, one with biallelic pLoF genotypes alone, another with biallelic pLoF and pDM genotypes, and the third with biallelic synonymous genotypes as negative control. Using REGENIE^27^, we performed recessive gene-based test with biallelic gene burdens with at least 4 biallelic carriers for 54 quantitative and 439 binary traits (**Supplementary Table 6**).

We found 13 significant associations under the recessive model (p<2.89×10^−7^, FDR~7.14%, **Supplementary Methods; Supplementary Figure 8B**; **Supplementary Table 14**), many of which had greater p-values and effect sizes under the recessive model than under the additive model (**Supplementary Figure 9**). We tested for dominance deviation by jointly modelling additive and dominant effects (**Methods**) to identify associations likely mediated by non-additive effects. Three associations had significant dominance deviation (p_domdev_<0.05/13=0.0038) with a clear recessive phenotypic pattern (**Figure 3G and 3H**). None of these had prior associations linking the gene to the phenotype:

- *NLRP10* and viral pneumonia (p_rec_ =6.11×10^−8^; p_domdev_ =0.0028) (**Figure 3G**), plus a suggestive association with “viral agents causing diseases classified elsewhere” (p_rec_ =4.01×10^−6^; p_domdev_=0.0028). These associations are consistent with a key role of *NLRP10* in the inflammasome pathway^57^.
- *HSD17B14* and disorders of bone density (p_rec_ =7.12×10^−8^; p_domdev_ =2.0×10^−5^) (**Figure 3G**), which is involved in steroid hormone metabolism^58^ and could thus have an indirect impact on bone health.
- *NCAPD2* and vitamin B12 and (p_rec_ =2.60×10^−7^; p_domdev_ =2.48×10^−4^) (**Figure 3H**). This gene encodes a condensin I complex subunit essential for chromosome condensation^59^ and additional studies are needed to establish its relevance to B12 metabolism.

More details on these associations and additional associations with suggestive p-values (p_rec_<5.0×10^−6^) are provided in **Supplementary Note 4** and **Supplementary Table 14**. Overall, these results suggest that increasing the sample size for recessive testing via meta-analysis across cohorts may yield further novel findings which have been missed by additive association testing, particularly if they involve very rare variants.

### Insights from human knockouts

#### Discovery of 2,991 genes with putative human knockouts

G&H exomes, as expected, showed higher rate of accrual of genes with one or more homozygous pLoF-HC genotypes compared to non-South Asian ancestry exomes in UK Biobank (**Figure 4A,B**). In the 44,028 G&H exomes, we identified a total of 2,991 genes with biallelic pLoF genotypes, referred to as putative human knockouts (**Supplementary Table 15**): 2,951 genes with 8,144 homozygous pLoF genotypes and 249 genes with 473 compound heterozygous pLoF genotypes. Genes with biallelic loss in G&H were depleted among the genes that are essential in cell culture or knockout lethal in mice and among the autosomal recessive disease genes (**Supplementary Note 6; Supplementary Figure 10; Supplementary Table 16**).

We compared the list of genes with human knockouts in G&H to those found in other genomic datasets, although note that these studies may not have as detailed phenotype information as G&H. Aggregating across 5 datasets (gnomAD^22^ v4 exomes which includes UK Biobank, RGC-ME^60^, deCODE^18^, PROMIS^19^, BiB^16^, there were 7,348 genes with human knockouts in approximately over 1.4 million individuals. Despite much smaller sample size, we found 546 additional genes that uniquely have human knockouts in G&H (**Figure 4C left**). We also found that 1,669 genes have human knockouts only in G&H compared to 2,778 genes with human knockouts in the 469,814 exomes from UK Biobank^61^, the only other cohort with broadly accessible phenotypic information (**Figure 4C right**).

In G&H, unlike in UK Biobank, the number of genes with putative human knockouts is growing close to linearly at the current sample size (**Figure 4D**), suggesting that sequencing of additional individuals will identify more genes with biallelic loss. Since the average level of autozygosity is higher among British-Pakistanis compared to British-Bangladeshis^17^, sequencing British-Pakistanis is generally expected to find more homozygous pLoF genotypes. However, there is slightly reduced genetic diversity in Pakistanis compared to Bangladeshis resulting from the historic bottleneck events mentioned above^24^. We found that, when conditioned on the level of autozygosity and sample size, sequencing British-Bangladeshis can maximize the number of unique pLoF variants (and genes) with at least one homozygous genotype, while sequencing Pakistanis can maximize the number of pLoF variants with more than one homozygous genotype (**Supplementary Note 5; Supplementary Figure 11; Supplementary Methods**).

#### Clinical utility of human knockouts in variant interpretation for autosomal recessive disease genes

The high autozygosity and rich phenotypic information in G&H can facilitate the assessment of the clinical impact of variants in autosomal recessive (AR) disease genes. While pLoF variants in disease genes known to be caused by loss-of-function mechanism are often automatically classified as likely pathogenic, computational predictions may have errors^22^. We found 368 Mendelian disease genes with AR inheritance^62^ that have individuals homozygous for pLoF variants in G&H (**Supplementary Figure 4D; Supplementary Table 17**). Of these pLoF variants, 63% are unreported in ClinVar and 12% have uncertain significance or conflicting interpretations (VUS/CI). We inspected the health records of the homozygous carriers of these variants and identified several examples where they provided supporting evidence that the variant was indeed pathogenic (**Supplementary Table 18**). To highlight a few:

- A stop-gain variant in *LPL* (chr8-19955849-C-T, p.Gln262Ter) with conflicting interpretations in ClinVar. Loss of this gene causes lipoprotein lipase deficiency (OMIM 238600) characterized by significantly elevated serum triglycerides and ectopic lipid deposition. One homozygous carrier of this variant had significantly elevated serum triglycerides (lifetime median of 15.7 versus cohort median of 1.5mM, z-test p=6.98×10^−5^) (**Figure 4E**) from their early 30s, lowered HDL cholesterol (0.4 versus 1.2mM, z-test p=8.68×10^−7^), diagnostic codes for E78 (disorders of lipoprotein metabolism, prescriptions of Omacor tablets (omega-3-acid ethyl esters used for hypertriglyceridemia), and other related complications including K85 (acute pancreatitis), type 2 diabetes, and steatohepatitis.
- A frameshift variant in *ABCC2* (chr10-99818879-CCT-C, p.Leu788ValfsTer13) with conflicting interpretations in ClinVar. Loss of this gene is implicated in Dubin-Johnson syndrome (OMIM 237500) with a clinical symptom of chronic cholestatic jaundice. One homozygous carrier of this variant had persistently raised bilirubin (56 versus 7uM, z-test p=1.52×10^−6^) (**Figure 4E**) from their early 30s and diagnosis codes for E80 (disorders of porphyrin and bilirubin metabolism). This phenotypic profile is comparable to that observed in another homozygous carrier of a likely pathogenic splice donor variant (chr10-99792360-G-A) including consistently elevated serum bilirubin (65 versus to 7uM, z-test p=2.94×10^−7^) from their mid-20s and diagnoses codes for E80, K76 (other disorders of liver), and steatohepatitis.

In these examples, the quantitative phenotypes relevant to the disease displayed clear recessive patterns (**Figure 4E**), confirming the autosomal recessive nature and the need for homozygous carriers to assess the clinical impact of these variants. These results illustrate G&H as a unique resource to guide variant interpretation for AR disease genes based on the abundance of homozygous genotypes and medical records.

#### Insights into drug development from human knockouts

The presence of human knockouts without significant adverse health outcomes can suggest that complete lifelong absence of the gene is viable, indicating that therapeutic antagonism of the gene may also be safe and well tolerated^14,16,20^. Using drug information from Open Targets^63^, we examined the enrichment of drugs that have human knockouts in their target genes over those without for trial phase transitions (**Methods**). Among 3,324 drugs with antagonistic modes of action, drugs with human knockouts are 2.2-times more likely to transition past phase 1 (p=5.3×10^−5^), the primary focus of which is safety and tolerability (**Figure 4F; Supplementary Table 19**). This pattern was specific to phase 1 transition, drugs with antagonistic modes of action, and drugs with non-oncology indications (**Supplementary Figure 12A; Supplementary Table 19**). Comparable association was observed when the analysis was subset to drugs with only one target gene or when testing whether all target genes (compared to any target gene above) of the drug have human knockouts (**Supplementary Figure 12B; Supplementary Table 19**).

Phenotypic profiles in human knockouts can help inform the therapeutic effects anticipated when antagonizing a gene. We highlight some examples in G&H:

- SLC10A2, a bile acid transporter, is targeted by several small molecule inhibitors primarily to treat biliary diseases. Two individuals homozygous for a frameshift variant (chr13-103052648-CA-C, p.Trp186GlyfsTer23) have markedly reduced levels of LDL cholesterol (lifetime median of 50.3 and 36.4 compared to cohort median of 116mg/dL). This is consistent with the significant reduction in LDL cholesterol levels observed in clinical trials (maximum 13.4-27.0mg/dL reduction)^64–68^. Interestingly, we did not observe notable alteration in the LDL cholesterol levels among the heterozygous carriers (median of 114.1 compared to 116mg/dL among noncarriers), suggesting that close to complete loss of *SLC10A2* may be necessary to yield changes in cholesterol levels.
- APOC3 is targeted by several antisense oligonucleotide drugs to treat hypertriglyceridemia. Consistent with previous reports^19^, we also observed one individual homozygous for a stop-gain variant (chr11-116830637-C-T, p.Arg19Ter) with 62% lower triglyceride level (lifetime median of 49.56 compared to cohort median of 130.98mg/dL). This level is comparable to the range of maximal triglyceride reductions (44 to 77%) reported in clinical trials^69–71^.
- NYP5R, an orexigenic neuropeptide, is a target of a small molecule inhibitor investigated for obesity. We found one individual homozygous for a frameshift variant (chr4-163350636-ATG-A, p.Val125PhefsTer24). The carrier had a diagnosis code for obesity (E66) and modestly elevated weight (lifetime median of 75kg) and BMI (29.3kg/m^2^) compared to the median observed in the sex-and ancestry-matched subset (65kg and 27.4kg/m^2^) of G&H. This is consistent with mild late-onset obesity observed in *Npy5r* knockout mice^72–74^ and only a modest weight loss effect (−1.1kg compared to placebo) observed in the clinical trial^75^.

Phenotypic information on human knockouts can also allow evaluation of potential safety issues of therapeutically antagonizing a gene. Below are some examples from G&H:

- HSD17B13 is the target of RNA interference drugs to treat non-alcoholic liver diseases, identified based on genetic association^76^. It is part of the hydroxysteroid (17b) dehydrogenase superfamily involved in steroid metabolism, raising potential considerations for reproductive health. We found four (including one from additional sequencing) homozygous carriers of two frameshift variants (chr4-87318347-ATCTCT-A, p.Glu98AspfsTer14 and chr4-87313944-CG-C, p.Ala192AspfsTer14). All three female carriers had medical records indicating successful pregnancy(s) with one reporting having had four healthy pregnancies at a follow up research visit. This suggests that absence of *HSD17B13* may not have an effect on the reproductive potential or pregnancy in females. The remaining male carrier’s health record was unremarkable. Overall, this aligns with the lack of major health or reproductive issues associated with HSD17B13 loss, as reported in phase 1 trials (NCT04565717 and NCT04202354), which included male and female participants of reproductive age^79,80^.
- IGF1R is targeted by several small molecule inhibitor or antibody drugs for cancer indications. Hyperglycaemia was reported as an adverse drug reaction for two inhibitory antibody drugs while hypoglycaemia was reported as a reason for early termination of a trial that tested recombinant IGF1, a ligand of IGF1R (NCT00330668). We found one individual homozygous for a frameshift variant (chr15-98957361-CGA-C, p.Arg1343ThrfsTer30). Compared to the BMI-matched diabetic subset of G&H, the homozygous carrier had an earlier onset of type 2 diabetes (age 39 versus 45) and displayed markedly higher HbA1c levels (89 versus 47mmol/mol) despite being prescribed three glucose-lowering medications at maximal doses. HbA1c levels were only mildly elevated among 6 heterozygous pLoF carriers (median of 41 compared to 38mmol/mol among noncarriers) similarly to 1.1mmol/mol increase observed among the heterozygous carriers of rare damaging missense variants in UK Biobank^81^.

These results show that the presence of human knockouts and detailed review of their phenotypes can provide meaningful insights for drug development, enabling the assessment of efficacy and possible safety risks of therapeutically targeting a gene.

## Conclusions

Our study demonstrates the power of large-scale exome sequencing in a South Asian-ancestry cohort with high autozygosity to drive novel biological discoveries. We identify over 100 previously unreported gene-phenotype associations and more than 500 additional genes with homozygous pLoF genotypes. We highlight valuable insights that can be gained through the phenotypic reviews of these “human knockouts”. As G&H sequences beyond 100,000 individuals, we anticipate identifying many additional human knockouts given the near linear growth observed thus far^82^.

Another valuable capability in G&H is the ability to recontact individuals for detailed characterization to enable mechanistic insights^14,15^. While profound advances can be made even with a single knockout individual in the presence of strong scientific priors^14–16^, recalling first-degree relatives of index knockout individuals can be an effective way to further solidify findings^19,20^ (**Supplementary Figure 11D**). Beyond the rich genomic and medical data currently presented, ongoing initiatives for detailed molecular phenotyping in G&H - encompassing transcriptomics, proteomics, and metabolomics - will further enhance the interpretability of genotype-phenotype relationships for these rare genotypes and expand functional allelic series beyond pLoF variants.

Overall, the G&H exome resource presents a valuable opportunity to advance biomedical research within the South Asian community which has been historically underrepresented in genomic research, while also broadening our understanding of and expanding therapeutic options for human health and disease.

## Methods

### Cohort description

Genes & Health is a longitudinal population genomic medicine study of currently over 65,000 individuals of South Asian ancestry living in the United Kingdom, with ongoing recruitment and cohort size target of >100,000 participants^47^. Adult volunteers aged 16 and over from self-reported British Bangladeshi and British Pakistani ethnicities have been recruited since 2015. At recruitment, each volunteer completes a brief questionnaire, provides an Oragene saliva sample (for DNA and genetic data), and consents for linkage to their longitudinal health records. This includes local primary (general practitioner or family doctor) and secondary (hospital) care electronic health records from UK National Health Service (NHS) alongside national datasets from NHS England, which contain Office for National Statistics mortality data (death registry with ICD10 coded cause of death), Hospital Episode Statistics Data (ICD10 coded inpatient and emergency department diagnoses), Cancer registry, among others. The demographic characteristics of the cohort have been previously described^47^. The current exome sequence dataset includes 44,028 individuals consisting of 59% British-Bangladeshi, 40% British-Pakistani, and other (1%) South Asian ancestry, and 56% females and 44% males. The work was conducted with an approval from the London South East NRES Committee of the UK Health Research Authority (14/LO/1240).

### Genetic and phenotypic data preparation

The **Supplementary Methods** contain details on the generation and quality control of exome sequencing data, statistical phasing, and the extraction and preparation of phenotypes. Supplementary Tables 20-23 show various details of the variant and genotype quality control.

### Variant annotation

Variants were annotated using the Ensembl Variant Effect Predictor (VEP v105) with LOFTEE plugin (v1.04_GRCh38)^22^. For all analyses, we used the predicted effect on the MANE transcript^21^. Predicted loss-of-function variants (pLoF) included frameshift, stop-gain, splice acceptor and donor variants and were further annotated by LOFTEE to be high- or low-confidence (pLoF-HC or pLoF-LC, respectively). Predicted damaging missense variants (pDM) were defined as CADD>20, Polyphen2>0.445, and SIFT deleterious. We used gnomAD (v4.1) to check for the presence of G&H variants in gnomAD or in the non-Finnish European (NFE) subset of gnomAD, and to compare allele frequency in G&H against that in gnomAD NFE subset.

### Clinical variation

We used ClinVar database^83^ (accessed November 13^th^, 2022) to annotate disease-relevant variants in G&H exomes. We analyzed variants labeled as pathogenic, likely pathogenic, or pathogenic/likely pathogenic in ClinVar (PLP), variants with uncertain significance (VUS), and variants with conflicting interpretations (CI). Disease inheritance pattern was obtained from the Online Mendelian Inheritance in Man (OMIM) database^62^ (accessed April 29^th^, 2024). The list of clinically actionable genes was derived from ACMG^84^ v3.2. For the comparison of ClinVar annotation against UK Biobank, we down-sampled the European-ancestry exomes from UK Biobank to match the sample size of G&H.

### Fine-scale population structure and founder variants

We defined a set of 17,172 unrelated G&H individuals (9,063 British-Bangladeshis and 8,109 British-Pakistanis) by removing one individual from each pair if they were related up to 3^rd^ degree and shared IBD segments >40cM. We used principal component analysis to explore population structure in G&H, combining the cohort with a reference panel composed of South Asian individuals from other cohorts (**Supplementary Figure 1**). To explore fine-scale structure within the British-Pakistanis, we used IBIS^85^ to infer identity-by-descent (IBD) regions and then clustered individuals based on their total IBD with other individuals using the Louvain method. We used Fisher’s exact tests to identify variants with significantly different frequencies between British-Pakistani sub-populations (**Supplementary Figure 2, Supplementary Tables 1 and 2**), considering only the variants that were sufficiently common to see a significant difference after multiple testing given the size of each cluster. Full details are provided in the **Supplementary Methods**.

### Rare variant association analyses under the additive model

We included 55 quantitative traits with measurements in at least 5,000 participants and 591 binary traits with at least 100 cases for rare variant association analyses. We used REGENIE^27^ v3 to carry out single variant and gene-based tests adjusting for age, sex, age^2^, and the first 20 PCs. For whole genome regression in step 1, we used variants from the exome sequence data with minor allele frequency (MAF)>1%, minor allele count (MAC)>100, missing call rate<10% and Hardy-Weinberg equilibrium (HWE) p-value<1×10^−15^ and samples with missing rate<10%. We also pruned the variants with PLINK using window sizes of 500 variants, shifted by 50 variants, and LD>0.2. In step 2, we ran single variant test for variants with MAC>=5 and gene-based tests (burden, SKAT, and SKAT-O) for four variant consequence masks (Mask A: pLoF-HC; Mask B: pLoF-HC and pDM; Mask C: all pLoF and missense; Mask D: synonymous) and four allele frequency cutoffs (singletons, <0.01%, <0.1%, <1%). This included 122,690 high-confidence predicted loss-of-function (pLoF-HC) variants, 14,958 low-confidence pLoF variants (pLoF-LC), 524,289 predicted damaging missense (pDM) variants, 1,179,722 other missense variants, and 805,502 synonymous variants (SpliceAI <0.1). The number of variants per mask ranged from 66,841 singleton variants in MASK A, up to 1,816,278 variants with AF <0.01 in MASK C (**Supplementary Table 24**). We applied a Firth correction for associations with p-values<0.01.

Genomic inflation factors were calculated as the median of the observed chi-squared test statistics divided by the expected median of the corresponding chi-squared distribution for each test and each phenotype (**Supplementary Note 3; Supplementary Table 7**). We used permutation to determine p-value cutoffs corresponding to a 5% false discovery rate (**Supplementary Methods**). Associations driven by synonymous variants and masks, variants in olfactory genes (HGNC “Olfactory receptors (OR)”) or the major histocompatibility complex (MHC, chr6:28510120-33480577), and associations that were not conditionally independent from nearby GWAS associations (**Supplementary Methods**) were omitted from the reporting of the number of associations.

Since binary traits were derived from ICD10 codes and custom code lists, there was some overlap in phenotype definitions, each with varying sensitivity and specificity. To accurately report the number of unique associations, we merged phenotypes with >75% reciprocal case overlap and applied text matching and manual curation to harmonize trait labels (**Supplementary Table 6**). All matches were manually reviewed. In total, 675 binary traits were consolidated into 591 unique phenotypes which were used to report the number of unique associations. Supplementary Tables contain results for all 675 binary traits.

We used a custom pipeline to annotate gene-phenotype pairs as “novel” or “reported” compared to GWAS/RVAS studies captured in Open Targets^63^ and Genebass^30^, and Mendelian diseases reported in OMIM^62^ (**Supplementary Methods**).

### Meta-analysis with UK Biobank

The analyses in UK Biobank included 425,618 individuals of European ancestry following the quality control measures previously described^30^, including heterozygosity and missing rates, sex concordance, and sex chromosome aneuploidy. We conducted rare variant association analyses for 33 cardiometabolic traits, including 13 binary traits and 20 quantitative traits (**Supplementary Table 6**), adjusting for age, sex, age^2^, age:sex, age^2^:sex, exome sequencing batch, and top 10 PCs. Comparable approaches to the analyses in G&H were used for whole genome regression (except for the use of chip genotype data) in step1 and association testing (including matching mask definitions) in step2 of REGENIE. Single variant and gene-based results from UK Biobank and G&H were meta-analyzed with a fixed-effects model weighted by the inverse of standard error using METAL. Genomic control was applied to adjust for population stratification. Results with inconsistent effect directions between UKB and G&H were filtered out.

### Recessive burden analyses

We performed statistical phasing for pLoF and pDM variants with MAF<5% following a previously described approach^56^ to identify compound heterozygous genotypes with high confidence (phasing probability>0.9) (**Supplementary Methods**; **Supplementary Figure 8A**). We then aggregated biallelic (homozygous or compound heterozygous) genotypes into recessive burden genotypes per gene (1 if the individual carried a biallelic genotype or 0 otherwise). Recessive gene burdens with at least 4 carriers were tested for association with 54 quantitative phenotypes (with measurements in at least 5,000 individuals) and 439 ICD10 codes (with at least 120 cases) (**Supplementary Table 6**), adjusting for age, sex, age:sex, age^2^, age^2^:sex, and top 20 PCs. We performed whole genome regression in step 1 using the variants filtered by the same parameters as in the additive analyses but in the subset of individuals with corresponding chip genotype data, then tested association with recessive genotype burden in step 2 of REGENIE. We also tested association with additive genotype burden for comparison. We used permutations to derive the p-value threshold of significance at p_rec_<2.89×10^−7^ corresponding to an FDR of 7.14% (**Supplementary Methods**; **Supplementary Figure 8B**). We additionally considered suggestive associations with p-values (p_rec_ <5.0 × 10^−6^).

Next, we investigated which of the identified associations have significant dominance deviation, using a 2-degrees of freedom test^13^. Specifically, we examined the following model to jointly test additive and non-additive effects:

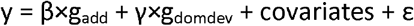

where g_add_ is encoded as [0,1,2] to represent additive genotypes and g_domdev_ is encoded as [0,1,0] to represent dominance genotypes. We used linear or logistic regression for quantitative or binary traits, respectively, using the same covariates among a subset of unrelated individuals. Due to the requirement of using unrelated individuals, the dominance deviation test could be underpowered compared to the mixed-model test by REGENIE which included related individuals. The significance p-value threshold was determined by adjusting for the number of associations considered for dominance deviation tests (13 for significant associations and 35 for suggestive associations).

### Identification and analyses of human knockouts

Human knockouts are defined as the homozygous or compound heterozygous carriers of pLoF-HC (MAF<1%) variants within the gene. For the comparison of accrual rate of genes with human knockouts across ancestry groups, we used exome sequence data from 469,815 participants of UK Biobank^61^ with the ancestry label previously reported^86^. Exome data in each ancestry group were down-sampled to match the maximum sample sizes available in different ancestry groups. For the comparison of genes with human knockouts from other genomic datasets, we used the list of genes either reported in previous publications^16,18,19^ (note that the criteria used to define human knockouts differ slightly depending on the cohort) or generated based on the variant-level information publicly available in gnomAD^22^ (v4.1) using the same criteria used in G&H. The comparison of pLoF genotype distributions between G&H British-Pakistanis and British-Bangladeshis is described in the **Supplementary Methods**. Details of the gene set enrichment analyses are available in **Supplementary Methods**.

### Human knockouts and drug phase transition

Drug target, indication, mode of action, and trial phase information was obtained from Open Targets^63^ drug dataset. For each drug, we derived the maximal trial phase and whether any or all of its target genes have human knockouts in G&H. Logistic regression test, along with Fisher’s exact test, was used to test the association between whether a drug has human knockouts and whether it passed certain trial phase adjusting for the number of target genes. To assess sensitivity, the analyses were repeated stratifying by antagonizing versus agonizing modes of action and oncology versus non-oncology indications, and among the drugs with only one target gene. Drugs listed as inhibitor, antagonist, blocker, negative allosteric modulator, antisense inhibitor, RNAi inhibitor, inverse agonist, disrupting agent, negative modulator, degrader, and allosteric antagonist were included as antagonistic drugs while those listed as activator, agonist, partial agonist, positive allosteric modulator, positive modulator, and stabilizer were included as agonistic drugs. Oncology indications were identified by performing a text match for the following terms: “cancer”, “neoplasia”, “neoplasm”, “leukemia”, “tumor”, or words ending in “oma”.

### Phenotypic review of homozygous carriers of pLoF variants in autosomal recessive disease genes for clinical interpretation

We focused on pLoF and pDM variants that are unreported or have unknown significance or conflicting interpretations in ClinVar which have homozygous carriers in G&H. For statistical testing on quantitative traits, we used z-test (in the case of one homozygous individual) or Wilcoxon rank sum test on log transformed traits. We also manually inspected the health record data to examine longitudinal changes in the quantitative traits, diagnostic codes, and medication prescriptions relevant to the disease.

## Data Availability

Summary-level data from the G&H 44,028 exomes are publicly available through a google cloud bucket (gs://genesandhealth_publicdatasets/results_44k_ExWAS/). Individual-level data are available within a Secure Data Environment upon application to G&H and approval by the Executive Committee.

## Author contributions

Conception and scientific oversight: H.K., H.C.M., and D.A.VH. Exome sequence data generation and quality control: D.A.VH., I.P., T.H., N.G. Phenotype extraction and quality control: B.M.J., D.A.VH., J.M.D., K.E. Variant and population genetic analyses: K.W., H.M.C., D.A.VH. Rare variant association analyses and downstream analyses: C.D., K.W., D.A.VH, P.M-S., A.M.M., K.C. Meta-analysis and downstream analyses: C.L., S.V.M., C.D. Recessive burden analyses: G.K., F.H.P., D.S.P., H.M.C. Human knockout identification and downstream analyses: H.K., G.K., M.A.A. Medical record review: B.M.J., J.G., S.F., D.A.VH. Clinical variant analyses: K.K., H.K., G.D.A. Consortium leadership and supervision: D.A.VH., S.F., R.C.T., G.D.A., S.P., E.R.H., J.C.M., L.A., R.M.T., K.E., S.L., J.M.M.H., Y.J., E.B.F., H.K., M.R.M., D.D. Consortium operations: K.A.H. Consortium project management: M.S. Consortium informatics: V.I. Writing – original draft: H.K., C.D., K.W., G.K., C.L., S.V.M., K.K., B.M.J., H.C.M., D.A.VH. Writing – review and editing: all authors.

## Acknowledgements

Genes & Health is/has recently been core-funded by Wellcome (WT102627, WT210561), the Medical Research Council (UK) (M009017, MR/X009777/1, MR/X009920/1), Higher Education Funding Council for England Catalyst, Barts Charity (845/1796), Health Data Research UK (for London substantive site), and research delivery support from the NHS National Institute for Health Research Clinical Research Network (North Thames). We acknowledge the support of the National Institute for Health and Care Research Barts Biomedical Research Centre (NIHR203330); a delivery partnership of Barts Health NHS Trust, Queen Mary University of London, St George’s University Hospitals NHS Foundation Trust and St George’s University of London. Current members of the Genes & Health Research Team are listed in full in the Supplementary Data.

Exome sequencing and analyses are funded by the Genes and Health Industry Consortium 1 of AstraZeneca PLC, Bristol-Myers Squibb Company, GlaxoSmithKline Research and Development Limited, Maze Therapeutics Inc., Merck & Co., Inc., Novo Nordisk A/S, Pfizer Inc., Takeda Development Centre Americas Inc. Genes & Health is/has also recently been funded by Alnylam Pharmaceuticals, 5 Prime Life Sciences, Phaeron, and Genomics PLC.

This research was funded in part by Wellcome (grant no. 220540/Z/20/A, Wellcome Sanger Institute Quinquennial Review 2021–2026). S.O. acknowledges funding from Wellcome 226800/Z/22/Z and 314234/Z/24/Z and NIHR Cambridge Biomedical Research Centre NIHR203312. F.H.L. is supported by the Wellcome Trust (award 224894/Z/21/Z) and the Medical Sciences Doctoral Training Centre at the University of Oxford. D.S.P. is supported by a Wellcome Trust Investigator award (221782/Z/20/Z).

We thank Social Action for Health, Centre of The Cell, members of our Community Advisory Group, and staff who have recruited and collected data from volunteers. We thank the NIHR National Biosample Centre (UK Biocentre), the Social Genetic & Developmental Psychiatry Centre (King’s College London), Wellcome Sanger Institute, and Broad Institute for sample processing, genotyping, sequencing and variant annotation. This work uses data provided by patients and collected by the NHS as part of their care and support. This research utilised Queen Mary University of London’s Apocrita HPC facility, supported by QMUL Research-IT, http://doi.org/10.5281/zenodo.438045

We thank: Barts Health NHS Trust, NHS Clinical Commissioning Groups (City and Hackney, Waltham Forest, Tower Hamlets, Newham, Redbridge, Havering, Barking and Dagenham), East London NHS Foundation Trust, Bradford Teaching Hospitals NHS Foundation Trust, Public Health England (especially David Wyllie), Discovery Data Service/Endeavour Health Charitable Trust (especially David Stables), Voror Health Technologies Ltd (especially Sophie Don), NHS England (for what was NHS Digital) - for GDPR-compliant data sharing backed by individual written informed consent.

## Most of all we thank all the volunteers participating in Genes & Health

A favourable ethical opinion for the main Genes & Health research study was granted by NRES Committee London - South East (reference 14/LO/1240) on 16 Sept 2014. Queen Mary University of London is the Sponsor and Data Controller.

Analyses with UK Biobank data were conducted under application IDs 26041 and 50314.

For the purpose of open access, the authors have applied a CC-BY public copyright license to any author accepted manuscript version arising from this submission.

## References

1. Bycroft, C. et al. The UK Biobank resource with deep phenotyping and genomic data. Nature 562, 203–209 (2018).

2. All of Us Research Program Genomics, I. Genomic data in the All of Us Research Program. Nature 627, 340–346 (2024).

3. Gaziano, J.M. et al. Million Veteran Program: A mega-biobank to study genetic influences on health and disease. J Clin Epidemiol 70, 214–23 (2016).

4. Nagai, A. et al. Overview of the BioBank Japan Project: Study design and profile. J Epidemiol 27, S2–S8 (2017).

5. Kurki, M.I. et al. FinnGen provides genetic insights from a well-phenotyped isolated population. Nature 613, 508–518 (2023).

6. Martin, A.R. et al. Clinical use of current polygenic risk scores may exacerbate health disparities. Nat Genet 51, 584–591 (2019).

7. Sirugo, G., Williams, S.M. & Tishkoff, S.A. The Missing Diversity in Human Genetic Studies. Cell 177, 26–31 (2019).

8. Williamson, A. & Fatumo, S. Genomic diversity improves disease discovery for all. Science 385, 255–256 (2024).

9. Ziyatdinov, A. et al. Genotyping, sequencing and analysis of 140,000 adults from Mexico City. Nature 622, 784–793 (2023).

10. Locke, A.E. et al. Exome sequencing of Finnish isolates enhances rare-variant association power. Nature 572, 323–328 (2019).

11. Kim, H.I. et al. Characterization of Exome Variants and Their Metabolic Impact in 6,716 American Indians from the Southwest US. Am J Hum Genet 107, 251–264 (2020).

12. Jacobs, B.M. et al. Genetic architecture of routinely acquired blood tests in a British South Asian cohort. Nat Commun 15, 8929 (2024).

13. Heng, T.H. et al. Widespread recessive effects on common diseases in a cohort of 44,000 British Pakistanis and Bangladeshis with high autozygosity. (medRxiv, 2024).

14. McGregor, T.L. et al. Characterising a healthy adult with a rare HAO1 knockout to support a therapeutic strategy for primary hyperoxaluria. Elife 9(2020).

15. Lam, B.Y.H. et al. MC3R links nutritional state to childhood growth and the timing of puberty. Nature 599, 436–441 (2021).

16. Narasimhan, V.M. et al. Health and population effects of rare gene knockouts in adult humans with related parents. Science 352, 474–7 (2016).

17. Malawsky, D.S. et al. Influence of autozygosity on common disease risk across the phenotypic spectrum. Cell 186, 4514–4527 e14 (2023).

18. Sulem, P. et al. Identification of a large set of rare complete human knockouts. Nat Genet 47, 448–52 (2015).

19. Saleheen, D. et al. Human knockouts and phenotypic analysis in a cohort with a high rate of consanguinity. Nature 544, 235–239 (2017).

20. Gurtan, A.M. et al. Identification and characterization of human GDF15 knockouts. Nat Metab 6, 1913–1921 (2024).

21. Morales, J. et al. A joint NCBI and EMBL-EBI transcript set for clinical genomics and research. Nature 604, 310–315 (2022).

22. Karczewski, K.J. et al. The mutational constraint spectrum quantified from variation in 141,456 humans. Nature 581, 434–443 (2020).

23. Malawsky, D.S. University of Cambridge (2021).

24. Arciero, E. et al. Fine-scale population structure and demographic history of British Pakistanis. Nat Commun 12, 7189 (2021).

25. Landrum, M.J. et al. ClinVar: public archive of relationships among sequence variation and human phenotype. Nucleic Acids Res 42, D980–5 (2014).

26. Miller, D.T. et al. ACMG SF v3.0 list for reporting of secondary findings in clinical exome and genome sequencing: a policy statement of the American College of Medical Genetics and Genomics (ACMG). Genet Med 23, 1381–1390 (2021).

27. Mbatchou, J. et al. Computationally efficient whole-genome regression for quantitative and binary traits. Nat Genet 53, 1097–1103 (2021).

28. Piel, F.B., Steinberg, M.H. & Rees, D.C. Sickle Cell Disease. N Engl J Med 376, 1561–1573 (2017).

29. Taher, A.T., Musallam, K.M. & Cappellini, M.D. beta-Thalassemias. N Engl J Med 384, 727–743 (2021).

30. Karczewski, K.J. et al. Systematic single-variant and gene-based association testing of thousands of phenotypes in 394,841 UK Biobank exomes. Cell Genom 2, 100168 (2022).

31. Kowalski, M.H. et al. Use of >100,000 NHLBI Trans-Omics for Precision Medicine (TOPMed) Consortium whole genome sequences improves imputation quality and detection of rare variant associations in admixed African and Hispanic/Latino populations. PLoS Genet 15, e1008500 (2019).

32. Uda, M. et al. Genome-wide association study shows BCL11A associated with persistent fetal hemoglobin and amelioration of the phenotype of beta-thalassemia. Proc Natl Acad Sci U S A 105, 1620–5 (2008).

33. Hodonsky, C.J. et al. Genome-wide association study of red blood cell traits in Hispanics/Latinos: The Hispanic Community Health Study/Study of Latinos. PLoS Genet 13, e1006760 (2017).

34. Vuckovic, D. et al. The Polygenic and Monogenic Basis of Blood Traits and Diseases. Cell 182, 1214–1231 e11 (2020).

35. Ding, K. et al. Genetic variants that confer resistance to malaria are associated with red blood cell traits in African-Americans: an electronic medical record-based genome-wide association study. G3 (Bethesda) 3, 1061–8 (2013).

36. Gurdasani, D. et al. Uganda Genome Resource Enables Insights into Population History and Genomic Discovery in Africa. Cell 179, 984–1002 e36 (2019).

37. Graham, S.E. et al. The power of genetic diversity in genome-wide association studies of lipids. Nature 600, 675–679 (2021).

38. Cirulli, E.T. et al. Genome-wide rare variant analysis for thousands of phenotypes in over 70,000 exomes from two cohorts. Nat Commun 11, 542 (2020).

39. Sun, C. et al. ADAM15 regulates endothelial permeability and neutrophil migration via Src/ERK1/2 signalling. Cardiovasc Res 87, 348–55 (2010).

40. Lu, D. et al. Inhibition of airway smooth muscle adhesion and migration by the disintegrin domain of ADAM-15. Am J Respir Cell Mol Biol 37, 494–500 (2007).

41. Lu, D., Scully, M., Kakkar, V. & Lu, X. ADAM-15 disintegrin-like domain structure and function. Toxins (Basel) 2, 2411–27 (2010).

42. Helkkula, P. et al. Genome-wide association study of varicose veins identifies a protective missense variant in GJD3 enriched in the Finnish population. Commun Biol 6, 71 (2023).

43. Castro-Ferreira, R., Cardoso, R., Leite-Moreira, A. & Mansilha, A. The Role of Endothelial Dysfunction and Inflammation in Chronic Venous Disease. Ann Vasc Surg 46, 380–393 (2018).

44. Huan, T. et al. Identifying Novel Genes and Variants in Immune and Coagulation Pathways Associated with Macular Degeneration. Ophthalmol Sci 3, 100206 (2023).

45. Carrillo-Carrasco, N., Chandler, R.J. & Venditti, C.P. Combined methylmalonic acidemia and homocystinuria, cblC type. I. Clinical presentations, diagnosis and management. J Inherit Metab Dis 35, 91–102 (2012).

46. van Meurs, J.B. et al. Common genetic loci influencing plasma homocysteine concentrations and their effect on risk of coronary artery disease. Am J Clin Nutr 98, 668–76 (2013).

47. Finer, S. et al. Cohort Profile: East London Genes & Health (ELGH), a community-based population genomics and health study in British Bangladeshi and British Pakistani people. Int J Epidemiol 49, 20–21i (2020).

48. Pessente, G.D. et al. Effect of Occurrence of Lamin A/C (LMNA) Genetic Variants in a Cohort of 101 Consecutive Apparent “Lone AF” Patients: Results and Insights. Front Cardiovasc Med 9, 823717 (2022).

49. Maghsoudi, S., Shuaib, R., Van Bastelaere, B. & Dakshinamurti, S. Adenylyl cyclase isoforms 5 and 6 in the cardiovascular system: complex regulation and divergent roles. Front Pharmacol 15, 1370506 (2024).

50. Agolini, E. et al. Expanding the clinical and molecular spectrum of lethal congenital contracture syndrome 8 associated with biallelic variants of ADCY6. Clin Genet 97, 649–654 (2020).

51. Tan, Y., Chrysopoulou, M. & Rinschen, M.M. Integrative physiology of lysine metabolites. Physiol Genomics 55, 579–586 (2023).

52. Razquin, C. et al. Lysine pathway metabolites and the risk of type 2 diabetes and cardiovascular disease in the PREDIMED study: results from two case-cohort studies. Cardiovasc Diabetol 18, 151 (2019).

53. Wang, Q. et al. Rare variant contribution to human disease in 281,104 UK Biobank exomes. Nature 597, 527–532 (2021).

54. O’Connor, M.J. et al. Recessive Genome-Wide Meta-analysis Illuminates Genetic Architecture of Type 2 Diabetes. Diabetes 71, 554–565 (2022).

55. Palmer, D.S. et al. Analysis of genetic dominance in the UK Biobank. Science 379, 1341–1348 (2023).

56. Lassen, F.H. et al. Exome-wide evidence of compound heterozygous effects across common phenotypes in the UK Biobank. Cell Genom 4, 100602 (2024).

57. Martinon, F., Mayor, A. & Tschopp, J. The inflammasomes: guardians of the body. Annu Rev Immunol 27, 229–65 (2009).

58. Lukacik, P. et al. Structural and biochemical characterization of human orphan DHRS10 reveals a novel cytosolic enzyme with steroid dehydrogenase activity. Biochem J 402, 419–27 (2007).

59. Martin, C.A. et al. Mutations in genes encoding condensin complex proteins cause microcephaly through decatenation failure at mitosis. Genes Dev 30, 2158–2172 (2016).

60. Sun, K.Y. et al. A deep catalogue of protein-coding variation in 983,578 individuals. Nature 631, 583–592 (2024).

61. Backman, J.D. et al. Exome sequencing and analysis of 454,787 UK Biobank participants. Nature 599, 628–634 (2021).

62. Amberger, J.S., Bocchini, C.A., Schiettecatte, F., Scott, A.F. & Hamosh, A. OMIM.org: Online Mendelian Inheritance in Man (OMIM(R)), an online catalog of human genes and genetic disorders. Nucleic Acids Res 43, D789–98 (2015).

63. Buniello, A. et al. Open Targets Platform: facilitating therapeutic hypotheses building in drug discovery. Nucleic Acids Res 53, D1467–D1475 (2025).

64. Palmer, M., Jennings, L., Silberg, D.G., Bliss, C. & Martin, P. A randomised, double-blind, placebo-controlled phase 1 study of the safety, tolerability and pharmacodynamics of volixibat in overweight and obese but otherwise healthy adults: implications for treatment of non-alcoholic steatohepatitis. BMC Pharmacol Toxicol 19, 10 (2018).

65. Newsome, P.N. et al. Volixibat in adults with non-alcoholic steatohepatitis: 24-week interim analysis from a randomized, phase II study. J Hepatol 73, 231–240 (2020).

66. Matsuyama, M. et al. Effects of Elobixibat on Constipation and Lipid Metabolism in Patients With Moderate to End-Stage Chronic Kidney Disease. Front Med (Lausanne) 8, 780127 (2021).

67. Yoshinobu, S. et al. Effects of Elobixibat, an Inhibitor of Ileal Bile Acid Transporter, on Glucose and Lipid Metabolism: A Single-arm Pilot Study in Patients with T2DM. Clin Ther 44, 1418–1426 (2022).

68. Bowlus, C.L. et al. Safety, tolerability, and efficacy of maralixibat in adults with primary sclerosing cholangitis: Open-label pilot study. Hepatol Commun 7(2023).

69. Witztum, J.L. et al. Volanesorsen and Triglyceride Levels in Familial Chylomicronemia Syndrome. N Engl J Med 381, 531–542 (2019).

70. Stroes, E.S.G. et al. Olezarsen, Acute Pancreatitis, and Familial Chylomicronemia Syndrome. N Engl J Med 390, 1781–1792 (2024).

71. Gaudet, D. et al. Plozasiran (ARO-APOC3) for Severe Hypertriglyceridemia: The SHASTA-2 Randomized Clinical Trial. JAMA Cardiol 9, 620–630 (2024).

72. Marsh, D.J., Hollopeter, G., Kafer, K.E. & Palmiter, R.D. Role of the Y5 neuropeptide Y receptor in feeding and obesity. Nat Med 4, 718–21 (1998).

73. Pedrazzini, T. et al. Cardiovascular response, feeding behavior and locomotor activity in mice lacking the NPY Y1 receptor. Nat Med 4, 722–6 (1998).

74. Kushi, A. et al. Obesity and mild hyperinsulinemia found in neuropeptide Y-Y1 receptor-deficient mice. Proc Natl Acad Sci U S A 95, 15659–64 (1998).

75. Erondu, N. et al. Neuropeptide Y5 receptor antagonism does not induce clinically meaningful weight loss in overweight and obese adults. Cell Metab 4, 275–82 (2006).

76. Abul-Husn, N.S. et al. A Protein-Truncating HSD17B13 Variant and Protection from Chronic Liver Disease. N Engl J Med 378, 1096–1106 (2018).

77. Sinnott-Armstrong, N. et al. Genetics of 35 blood and urine biomarkers in the UK Biobank. Nat Genet 53, 185–194 (2021).

78. Ruth, K.S. et al. Using human genetics to understand the disease impacts of testosterone in men and women. Nat Med 26, 252–258 (2020).

79. Mak, L.Y. et al. A phase I/II study of ARO-HSD, an RNA interference therapeutic, for the treatment of non-alcoholic steatohepatitis. J Hepatol 78, 684–692 (2023).

80. Sanyal, A.J. et al. Phase 1 Study of the RNA Interference Therapeutic ALN-HSD in Healthy Adults and Patients with Nonalcoholic Steatohepatitis. (2023).

81. Gardner, E.J. et al. Damaging missense variants in IGF1R implicate a role for IGF-1 resistance in the etiology of type 2 diabetes. Cell Genom 2, None (2022).

82. Minikel, E.V. et al. Evaluating drug targets through human loss-of-function genetic variation. Nature 581, 459–464 (2020).

83. Landrum, M.J. et al. ClinVar: improvements to accessing data. Nucleic Acids Res 48, D835–D844 (2020).

84. Miller, D.T. et al. ACMG SF v3.2 list for reporting of secondary findings in clinical exome and genome sequencing: A policy statement of the American College of Medical Genetics and Genomics (ACMG). Genet Med 25, 100866 (2023).

85. Seidman, D.N. et al. Rapid, Phase-free Detection of Long Identity-by-Descent Segments Enables Effective Relationship Classification. Am J Hum Genet 106, 453–466 (2020).

86. Karczewski, K.J. et al. Pan-UK Biobank GWAS improves discovery, analysis of genetic architecture, and resolution into ancestry-enriched effects. (medRxiv, 2024).

