## Supplementary Data for "Exome sequencing and analysis of 44,028 British South Asians enriched for high autozygosity"

### Table of Contents

Supplementary Figures

Supplementary Notes

Supplementary Methods

Supplementary References

List of current members of the Genes & Health Research Team

### Supplementary Figures

#### Supplementary Figure 1

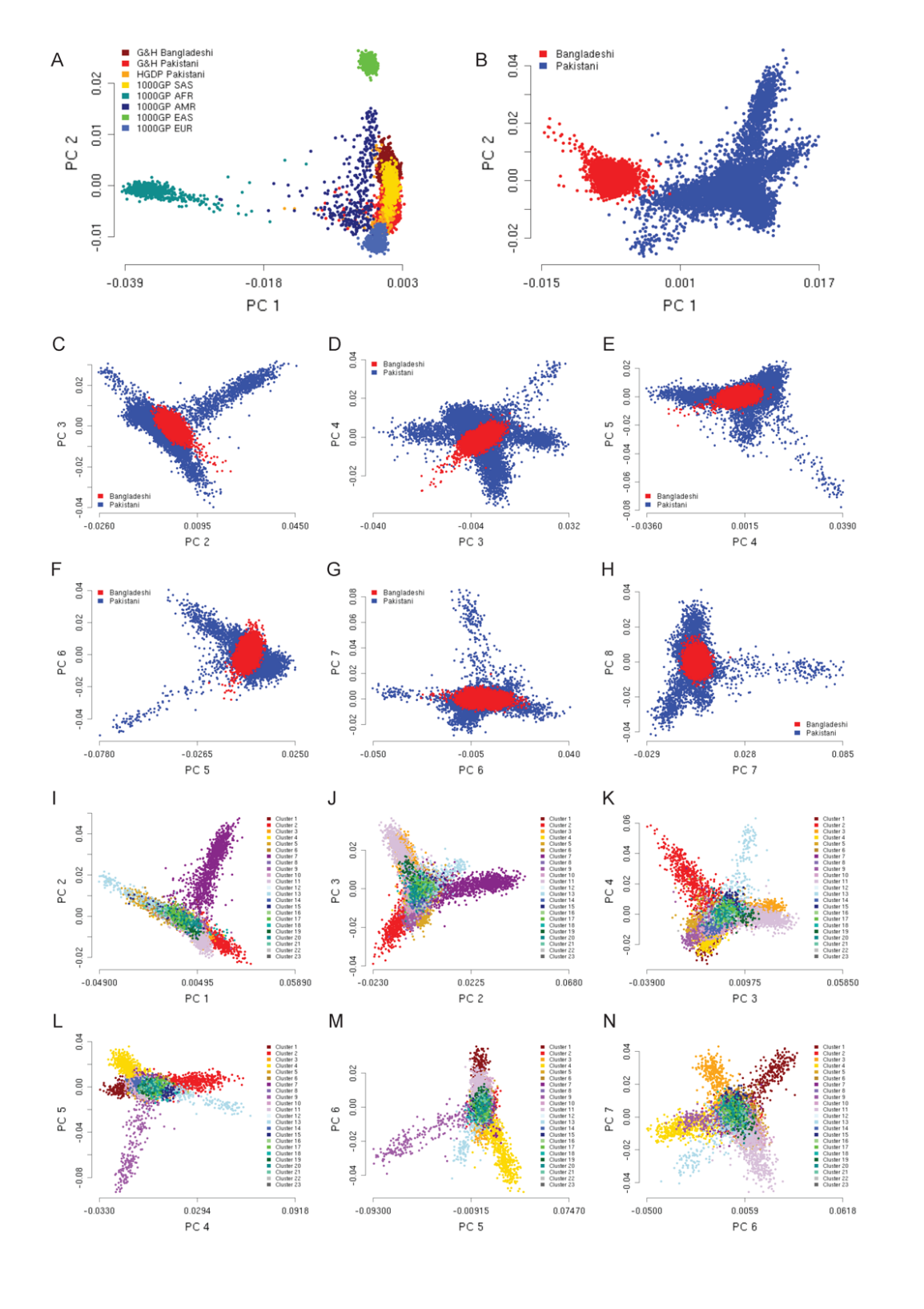

Principal component analysis (PCA) to survey population structure in G&H. (A-H) PCA results of G&H individuals together with reference populations. (A) PC1 against PC2 of G&H British Bangladeshis and Pakistanis together with a reference set comprising 1000GP and Pakistanis from HGDP. (B-H) PCA of G&H British Bangladeshis and Pakistanis using a South Asian reference set comprising South Asians from 1000GP and Pakistanis from HGDP and BiB. (I-N) PCA of G&H Pakistanis without a reference set and colored by cluster membership.

#### Supplementary Figure 2

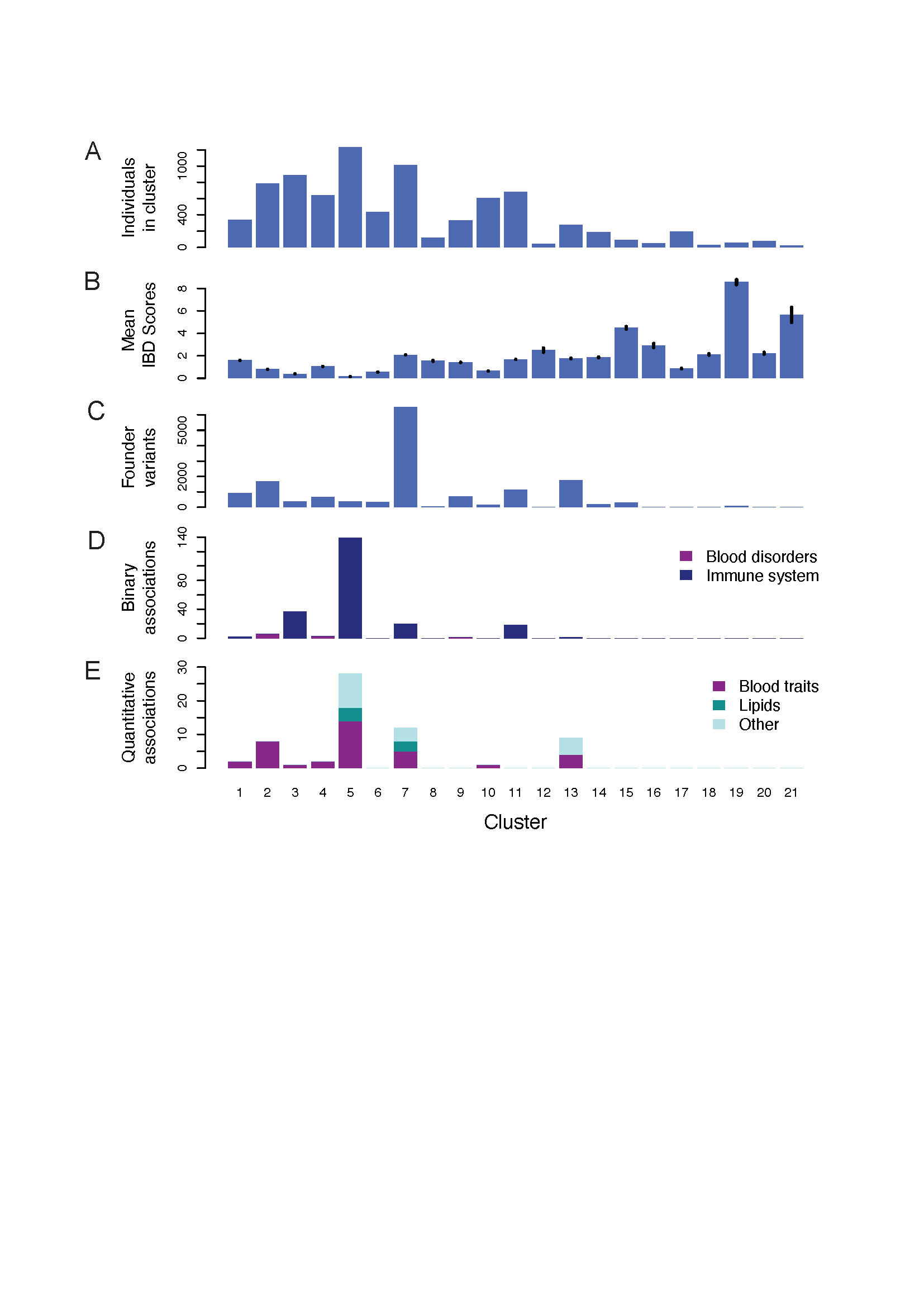

Sub-populations and putative founder effects in British Pakistanis from G&H. Panels (A-E) G&H British Pakistani population only. Metrics and association results for 21 population clusters. Clusters were generated by Louvain clustering of shared IBD segments of the combined unrelated G&H British Pakistanis and SAS reference populations from 1000GP, HGDP and BiB. (A) Number of unrelated G&H British Pakistanis in each cluster. (B) Mean IBD scores per cluster. These are the average total length of IBD segments shared between two individuals within a cluster. (C) Number of putative founder variants per cluster identified by Fisher's exact tests, where each cluster was tested against all other clusters combined. (D) Number of binary trait associations linked to putative founder variants in each cluster. (E) Number of quantitative traits associations linked to putative founder variants in each cluster. Numbers are shown in **Supplementary Table 1**.

#### Supplementary Figure 3

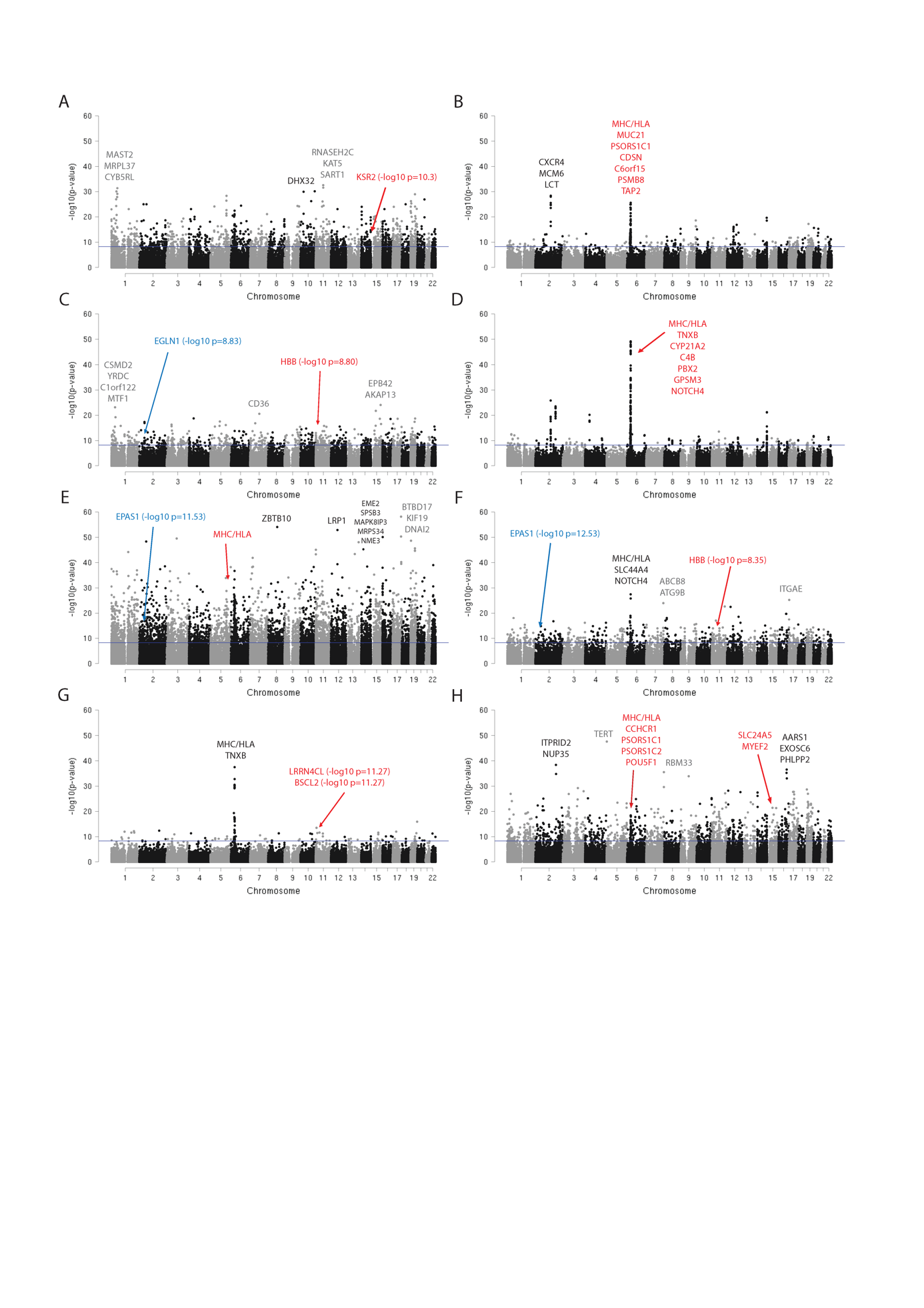

Manhattan plots show –log10 p-values for enrichment (Fisher's exact test) of putative founder variants by chromosomal location. Enrichment p-values are shown for the following clusters, (A) 2, (B) 3, (C) 4, (D) 5, (E) 7, (F) 9, (G) 10 and (H) 13. The blue line indicates the significance p-value threshold of p=5.25x10^-9^. Genes linked to the five most significant founder variants are labelled in black or grey, depending on the chromosomal color. Genes linked to significant phenotype associations are labelled in red, and genes found in the literature to be linked to altitude adaptation are labelled in blue. Some variants were linked to more than one gene.

#### Supplementary Figure 4

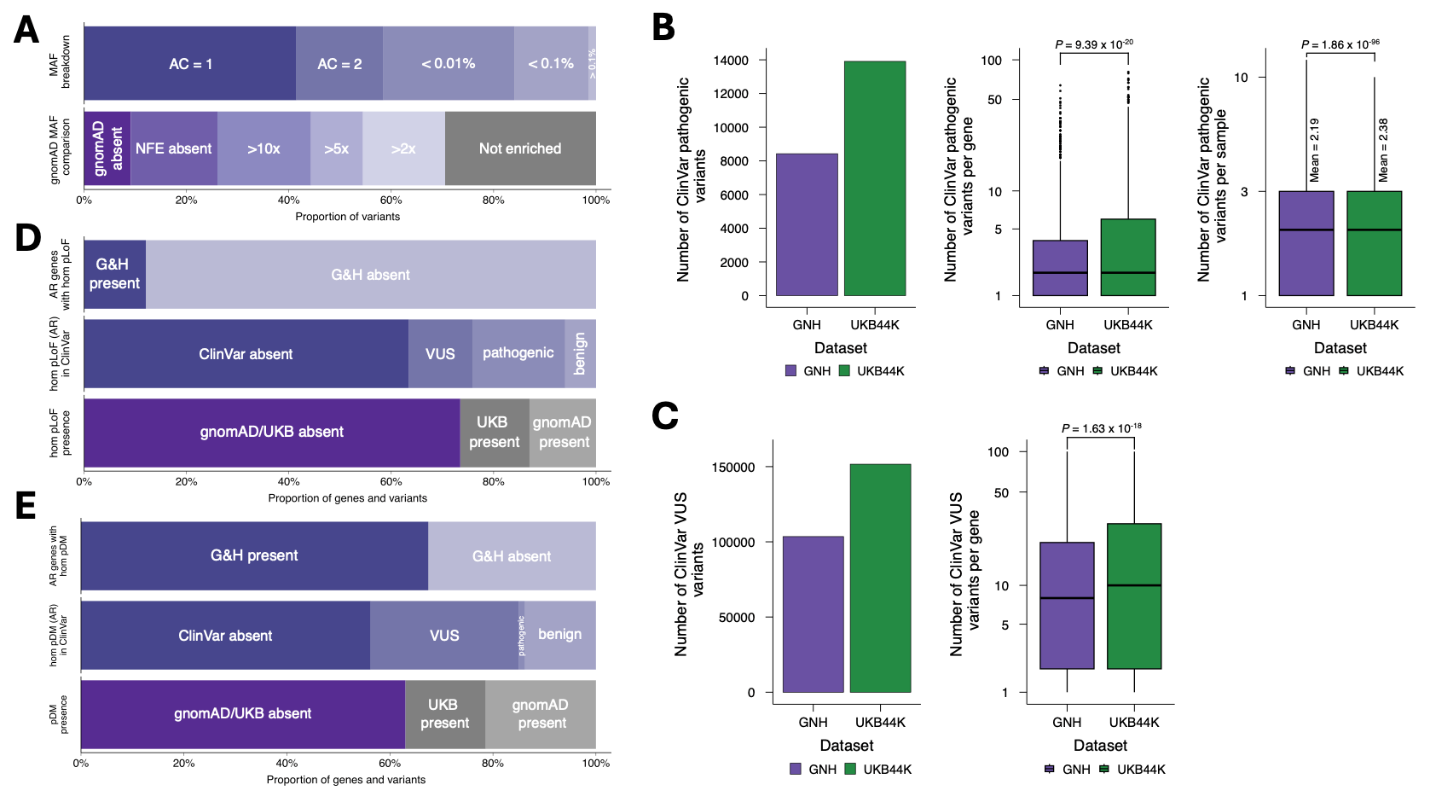
Survey of ClinVar variants and homozygous pLoF genotypes in AR disease genes in G&H exomes. (A) ClinVar Pathogenic variants in G&H stratified by allele count or frequency bins (upper) and by allele frequency comparison to gnomAD (lower). 10-fold increase (10x); 5-fold increase (5x); 2-fold increase (2x). (B) Comparison of the number of ClinVar PLP variants between G&H and UKB-EUR. Total number (left); number per gene (middle); number per individual (right). (C) Comparison of the number of VUS/CI variants between G&H and UKB-EUR. Total number (left); number per gene (middle); number per individual (right). Two-sample independent t-test was used for statistical testing. Upper and lower limits of the boxes: interquartile ranges; center lines: median; whiskers extend to values up to 1.5 times the interquartile range. (D) Proportion of AR disease genes with homozygous pLoF genotypes in G&H (top), ClinVar classification status of the pLoF variants (middle), presence of homozygous carriers of pLoF variants either absent or VUS/CI in ClinVar in UK Biobank or gnomAD (bottom). (E) Same as (D) for pDM variants.

#### Supplementary Figure 5

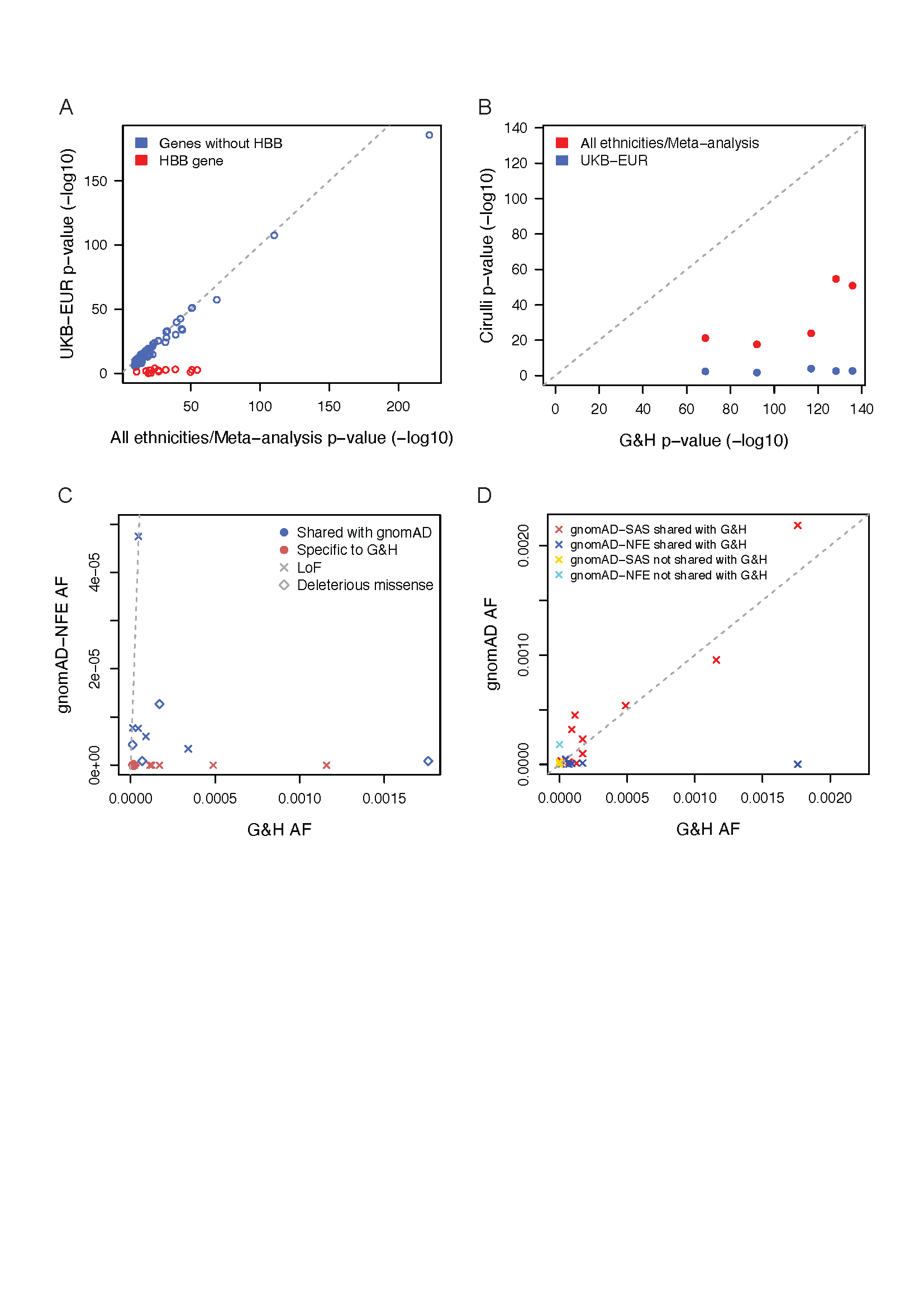

Comparison of *HBB* associations and variants between G&H and UKB. (A) Meta-analysis p-values for genes passing multiple-correction (p < 3.4e−10) across different ethnicities versus p-values from UKB-EUR using Cirulli *et al.^1^* Supplementary Data 2. Association p-values for all genes were highly correlated apart from the *HBB* gene. The -log10 p-values for the *HBB* gene, shown in red, are much decreased in UKB-EUR compared to the -log10 p-values from the meta-analysis across different ethnicities. (B) Phenotypes matched between G&H and Cirulli *et al.* that were associated with *HBB* show that even p-values from 44k G&H exomes are more significant than from the trans-ethnic meta-analysis (N~70k) in Cirulli *et al*. (C) Comparison of G&H AF with gnomAD NFE AF for fourteen LoF and eight deleterious missense variants that contributed to the gene-based associations in the *HBB* gene. All G&H AFs are larger or equal to gnomAD NFE AFs. Red points mark variants that are specific to G&H, i.e. they are absent in gnomAD NFE. (D) Comparison of gnomAD SAS and NFE LoF allele frequencies with G&H AFs in the *HBB* gene. LoF allele frequencies of gnomAD SAS are similar to G&H for shared variants, whereas LoF AF of gnomAD NFE are much lower than G&H AF. LoF AF for variants specific to gnomAD are very low, apart from one NFE stop gained variant, chr11-5226774-G-A with NFE AF = 0.00018.

#### Supplementary Figure 6

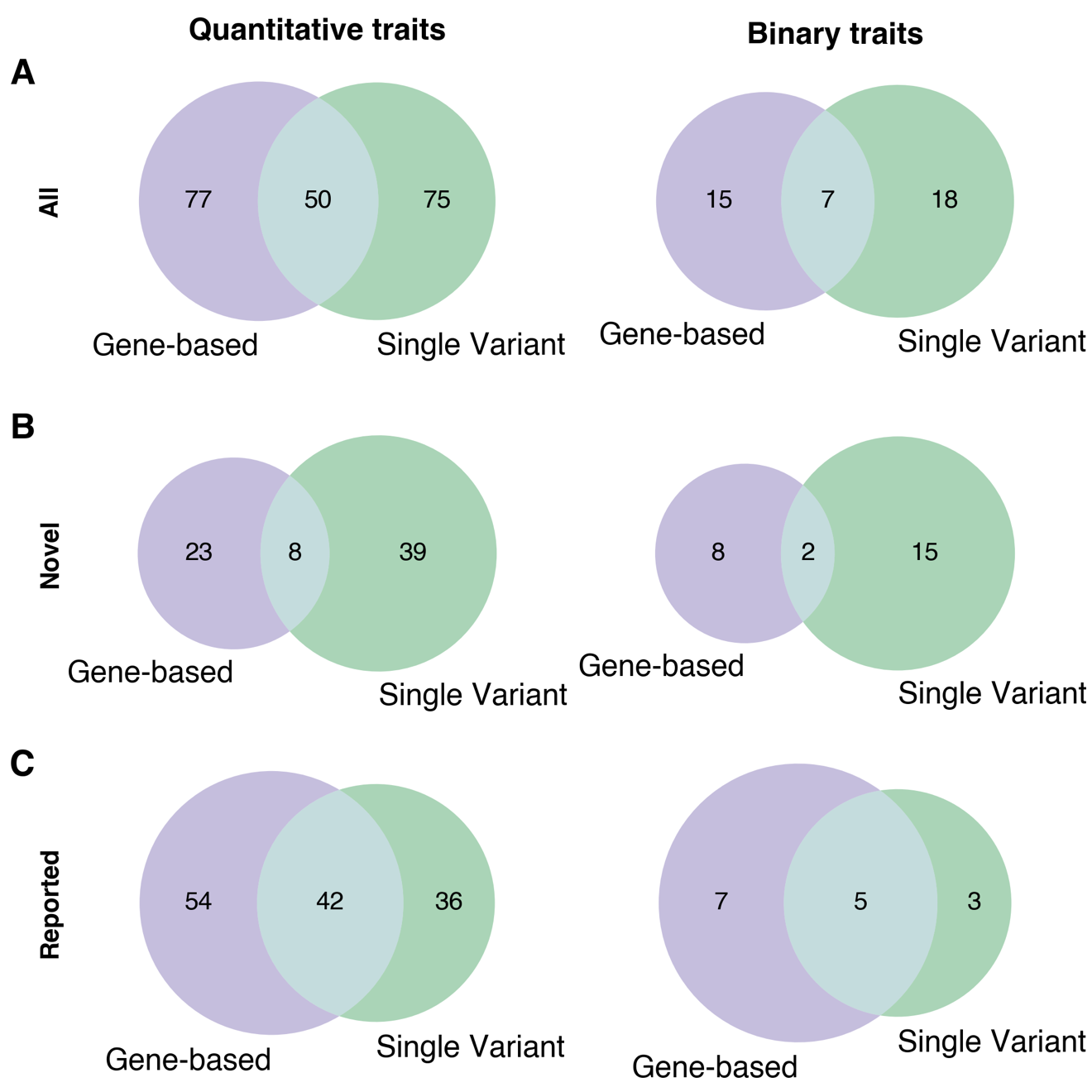

Counts of significant findings from additive exome-wide association studies. Number of unique gene-phenotype pairs implicated by gene-based (purple) or single variant predicted loss-of-function or predicted deleterious missense variants (green) for (A) all gene-phenotype pairs, (B) novel gene-phenotype pairs, and (C) reported gene-phenotype pairs.

#### Supplementary Figure 7

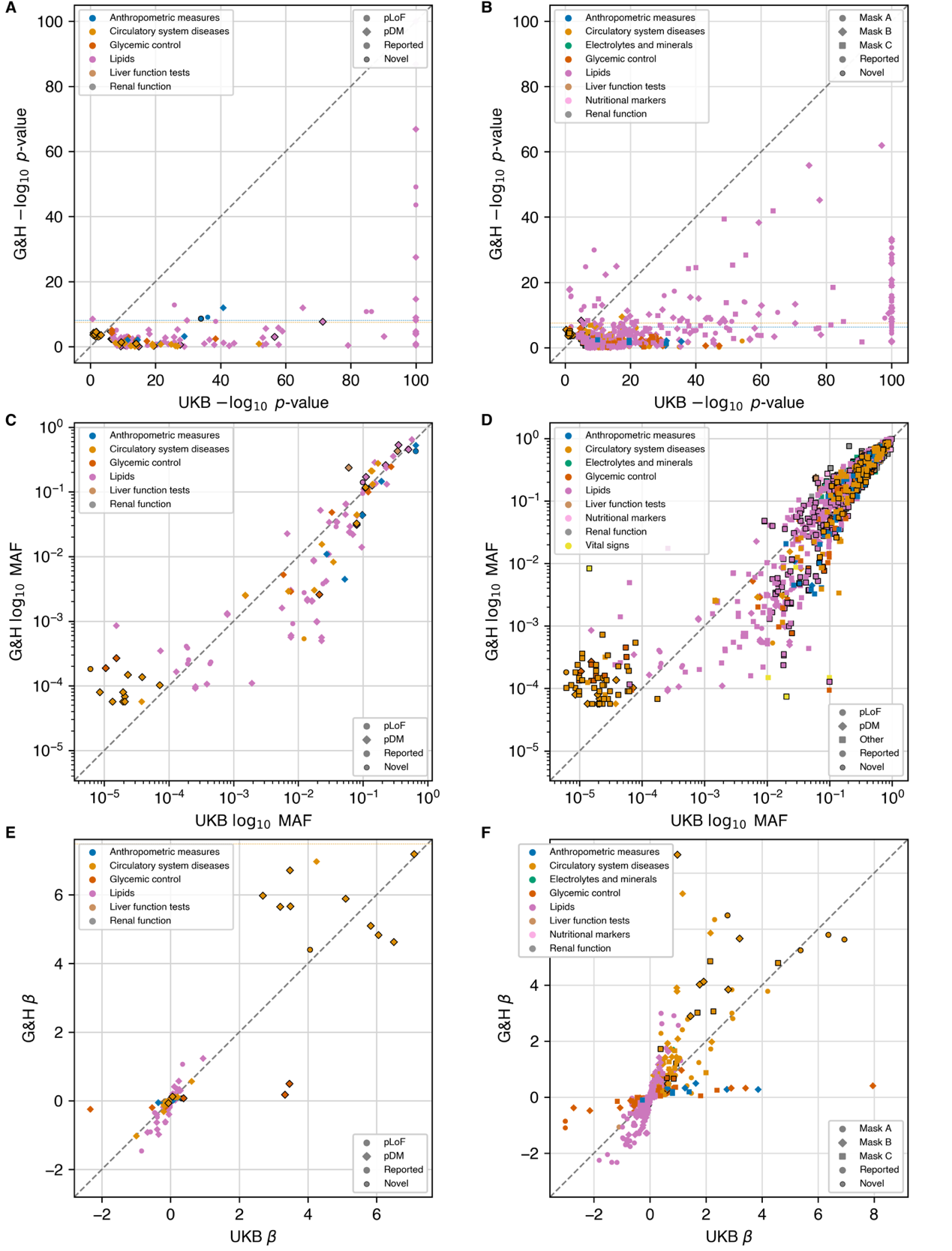

Comparison of p-values, allele frequency, and effect size between G&H and UKB for associations significant in the meta-analysis of cardiometabolic traits between these cohorts. G&H -log10 p-value vs. UKB -log10 p-value for (A) single variant pLoF or pDM associations and (B) gene-based associations. G&H MAF vs UKB MAF for (C) significant pLoF or pDM variants or (D) all significant variants. G&H effect size (beta) vs UKB effect size for (E) pLoF or pDM variants and (F) gene-based associations. All associations have been filtered to remove olfactory variants/genes; MHC variants/genes; and associations that are not conditionally independent of nearby GWAS signals.

#### Supplementary Figure 8

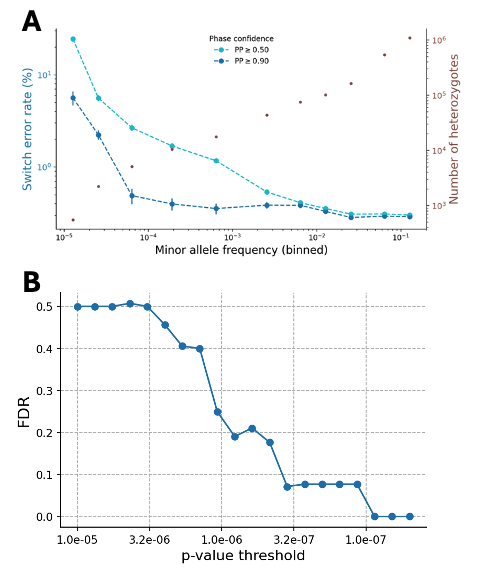

Data preparation for recessive gene-based association testing. (A) Switch error rates (SER) calculated based on 100 trios, using all available genotypes (PP>0.5; m=611,646), and those with high confidence (PP>0.9; m=606,011) (Methods). We also report the numbers of heterozygous genotypes (in brown) per frequency bin. PP: posterior probability. (B) False Discovery Rate (FDR) calculated for several thresholds, where false positives (FP) are determined using p-values from permutation-based tests (Methods), and true positives (TP) refer to significant associations identified in the original analysis. We note that the smallest FP was 9.77×10^-8^, resulting in FDR=0%, though we chose 2.89×10^-7^ as our study-wide significance threshold so the FDR here (7.14%) is closer to that of the main analysis (5%).

#### Supplementary Figure 9

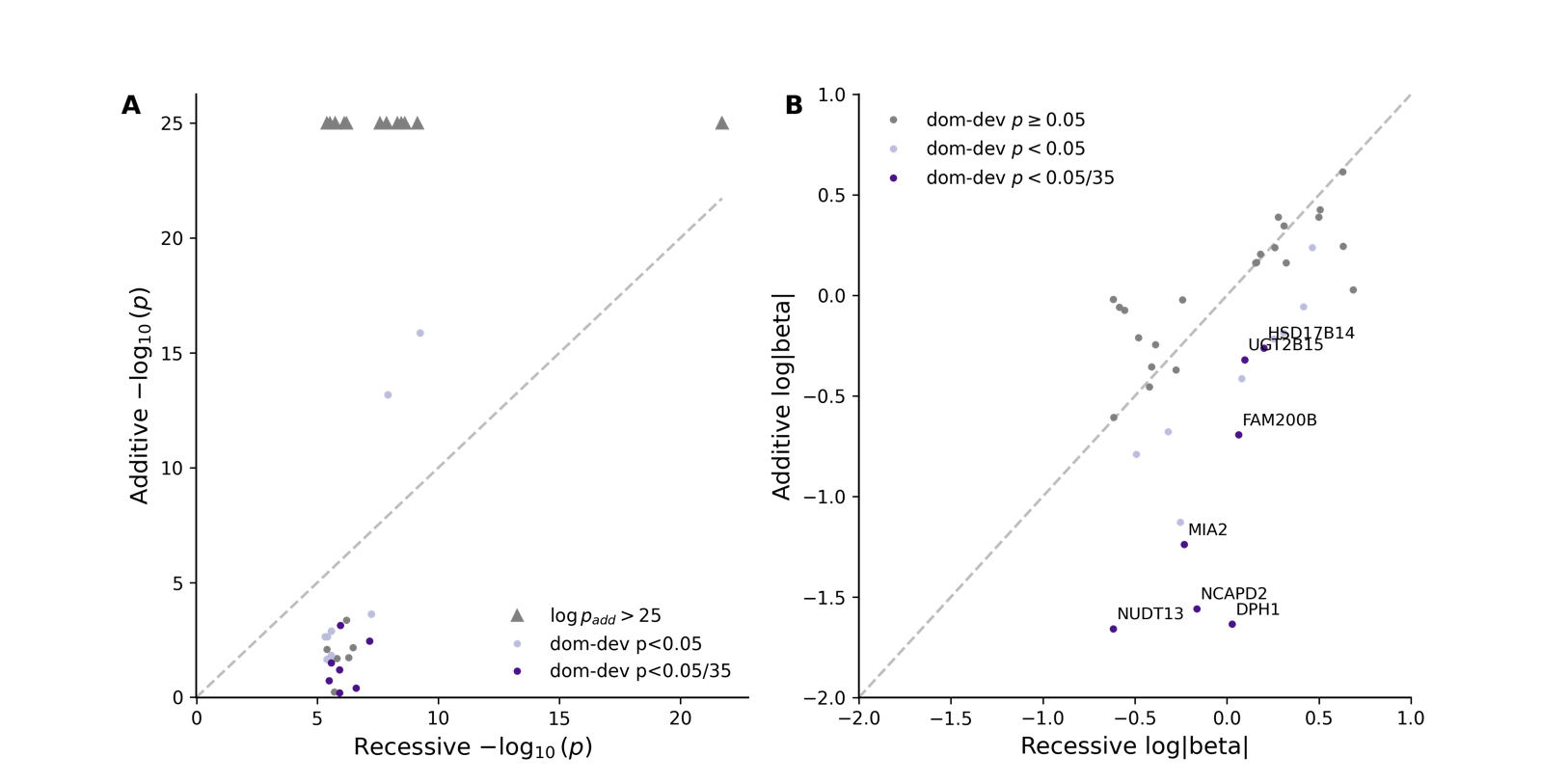

Comparison of recessive and additive effect sizes and p-values from the recessive burden analysis. (A) We contrast the recessive p-value to the one from the corresponding additive test, for all gene-trait associations having p_rec_<8.38×10^-8^. Associations with p_add_<5×10^-25^ are capped and denoted by triangles. The shading denotes the significance of the dominance deviation p-value (dom-dev p). (B) We contrast the recessive effect estimate (absolute value, log) to the one from the corresponding additive test, for all gene-trait associations having p_rec_<5.0×10^-6^ (suggestive associations; **Supplementary Table 14**). In both panels, we highlight with light blue those cases with nominal dominance deviation (p<0.05) and purple (also using labels in the right panel) for those with significant dominance deviation (p<0.05/35), accounting for the number of dominance tests performed; see Methods.

#### Supplementary Figure 10

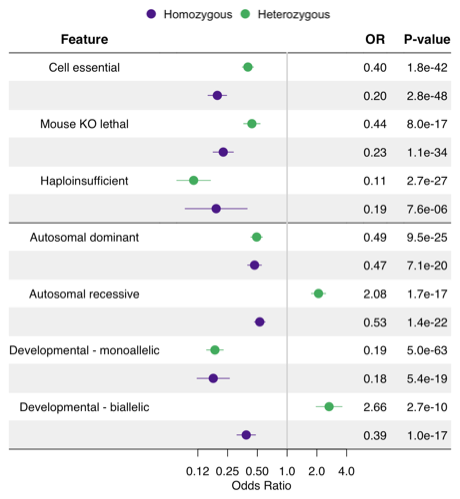

Enrichment of genes with heterozygous or homozygous pLoF genotypes in functional or disease gene sets. Logistic regression was used for statistical testing adjusting for relevant covariates.

#### Supplementary Figure 11

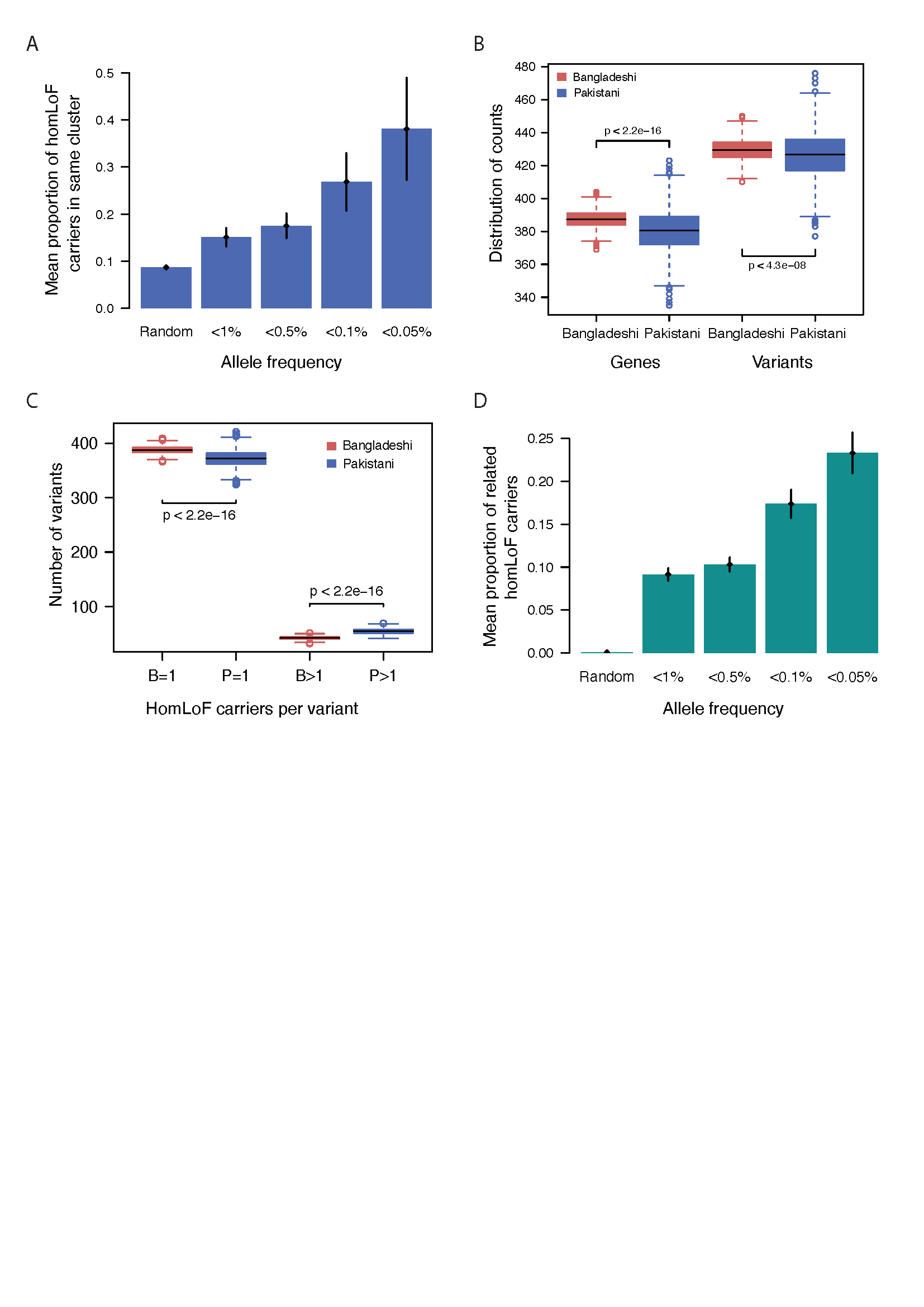

Implications of fine-scale population structure and relatedness in G&H for distribution and discovery of homozygous pLoF genotypes. (A) Proportion of 100,000 unrelated random pairs and mean proportion of unrelated Pakistani homozygous pLoF carriers who fall into the same population cluster at different allele frequency cutoffs. The plot shows that Pakistanis who share the same homozygous pLoF genotype are more likely to be from the same cluster at lower allele frequencies. (B) Distribution of number of variants and number of genes with homozygous pLoF genotypes after matching unrelated Bangladeshis and Pakistanis by FROH. P-values obtained from *t*-tests indicate a significantly lower number of genes and variants with homozygous pLoF genotypes in the Pakistanis. (C) Number of variants with either one homozygous carrier in Bangladeshis (B=1) and Pakistanis (P=1) or more than one homozygous carrier in Bangladeshis (B>1) and Pakistanis (P>1). Bangladeshis have significantly more genes and more variants with one carrier per gene and per variant than Pakistanis (*t-*test p<2.2x10^-16^), while Pakistanis have significantly more genes and more variants with more than one carrier per gene and per variant than Bangladeshis (*t*-test p<2.2x10^-16^). (D) Proportion of 100,000 random pairs and mean proportion of homozygous pLoF carriers with first-degree relationships at different allele frequency spectrums. People who share the same homozygous pLoF genotype are more likely to be close relatives at lower allele frequencies. In both panels the black lines indicate the standard error of the mean.

#### Supplementary Figure 12

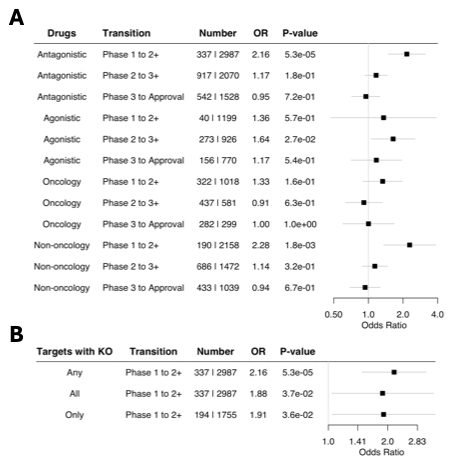

(A) Among the drugs with antagonistic mechanism of action, enrichment of drugs with human knockouts in their target genes versus those without per clinical trial status. (B) Sensitivity of the enrichment pattern depending on how drugs with human knockouts are defined, i.e., if any or all of the target genes have human knockouts or among the drugs with only one target gene. Logistic regression was used for statistical testing adjusting for relevant covariates.

### Supplementary Notes

#### Supplementary Note 1: Identification of putative founder variants in British Pakistanis and their association with traits

##### Identification of putative founder variants

To identify putative founder variants that have risen to higher frequencies in particular subgroups, we compared allele frequency (AF) between each cluster versus all others using Fisher’s exact test. We carried out power calculations to find the minimum allele count at which we would have power to detect a significant enrichment for a given cluster. Following that, we determined a Bonferroni-corrected p-value threshold of p<5.25x10^-9^ (**Methods**)**.** We identified 15,202 putative founder variants in 15,262 variant-cluster pairs with significant AF enrichment (**Supplementary Figures 2 and 3, Supplementary Table 2**) of which only 60 variants were significantly enriched in two different clusters, and 2,143 (14%) variants were only seen in a single cluster. The number of founder variants per cluster was predicted well by a model that included an interaction term between sample size and IBD score (r^2^=0.79; p=1.21x10^-6^, **Supplementary Table 3**).

The majority of the putative founder variants were either intronic (38%), missense (33%) or synonymous (22%), but 187 variants were high-confidence loss-of-function (pLoF-HC) and another 45 variants were low-confidence LoF (pLoF-LC). In total, 9,497 genes were affected by significant putative founder variants, and of those, 178 genes contained pLoF-HC variants, and 1,037 genes contained predicted deleterious missense variants with founder effects. 32 putative founder variants were either associated with OMIM phenotypes or pathogenic in ClinVar (**Supplementary Table 2**). Five percent of the founder variants are in the MHC/HLA region, including 38 of the 60 variants that were significantly enriched in two different clusters.

##### Association of putative founder variants with traits

We found that 139 putative founder variants were involved in 267 associations across 39 phenotypes. The associated binary phenotypes included sixteen immune traits (e.g. allergic and chronic rhinitis, ankylosing spondylitis, asthma, coeliac disease and vitiligo), as well as four blood disorders, such as thalassaemia and sickle-cell disorders (**Supplementary Figure 2D**). We also found associations with quantitative phenotypes, mainly red blood cell measurements, as well as lipid traits, bilirubin and vitamin B12 (**Supplementary Figure 2E**). The majority of these associations, 82%, were linked to the MHC/HLA region. It is plausible that some of these ‘founder variants’ rose to higher frequency in particular sub-populations due to historic positive selection incurred by environmental challenges in certain geographical locations^2^. For example, we identified two *HBB* variants with elevated frequencies in certain clusters, which accounted for 6 of the 104 significant single-variant associations. These variants may have risen to higher frequency in certain groups due to their role in conferring resistance to endemic malaria^3^ or possibly facilitating adaptation to high altitude^4^, rather than genetic drift. Several other genes reported to be under positive selection for high altitude adaptation, such as *EPAS1* and *EGLN1^5-8^,* also had variants enriched in certain clusters. Further work is required to determine the cause of the altered allele frequency in certain Pakistani sub-populations.

#### Supplementary Note 2: Survey of disease relevant and clinically actionable variants in G&H exomes

##### Survey of ClinVar variants

We found 8,450 variants curated as pathogenic or likely pathogenic (PLP) in ClinVar^9^ with at least one heterozygous or homozygous genotype in G&H. The majority were present at ultra-low frequency as expected (**Supplementary Figure 4A, Supplementary Table 4**). When compared to gnomAD, 3,745 PLP variants (44%) were absent, or were absent in the non-Finnish European (NFE) subset or have >10-fold higher allele frequency compared to that in gnomAD NFE (**Supplementary Figure 4A**). One notable example was the *HBB* gene which has 23 PLP variants in G&H. Of these, 12 were private to G&H, and 4 had significantly higher frequency in G&H compared to gnomAD NFE. Notably, a pathogenic missense variant (chr11_5226943_C_T, p.Glu27Lys), which is extremely rare in gnomAD NFE (AF=8.8x10^-6^), has a drastically elevated frequency in G&H (AF=0.016), and was significantly associated with 16 phenotypes, mainly red blood cell traits, thalassaemia, and sickle-cell disorder. Of the 104 significant associations by variants in *HBB* gene, involving 16 variants and 19 phenotypes, 92 (88%) were driven by 13 variants.

##### ACMG variants

Of particular importance are clinically actionable genes as defined by the American College of Medical Genetics and Genomics^10^ (ACMG SF v3.2). We found that 2,066 individuals (4.7%) are heterozygous or homozygous for at least one of 556 PLP variants across 81 ACMG genes. This included 1,012 individuals heterozygous for PLP variants in autosomal dominant genes and 7 individuals homozygous for PLP variants in autosomal recessive genes. It is important to note that the disease penetrance of the variants can vary considerably. There were 11 pLoF or pDM variants in four genes (*ATP7B, MUTYH, BTD,* and *GAA*) with significantly higher frequency in G&H compared to gnomAD NFE.

##### Comparison of ClinVar variants between G&H and UKB-EUR

It has been recognized that existing clinical variant databases may be biased towards European ancestry populations^11,12^. We compared the percentage of the variants that are present in ClinVar among the pLoF and pDM variants in G&H exomes versus those in a size-matched subset of European-ancestry exomes from UK Biobank (UKB-EUR). Among the variants that are uniquely found in each ancestry group, we found that a smaller portion of pLoF (3.5% versus 7.0%) and pDM (5.3% versus 6.1%) variants in G&H are present in ClinVar compared to UKB-EUR (**Supplementary Table 5**). This observation, along with another recent report specifically focused on variants in breast cancer genes^13^, suggests that there may be considerable under-representation of South Asian variants in ClinVar compared to European variants.

We next compared the classification status of the variants in ClinVar between G&H and UKB-EUR. We found greater portion of variants annotated as pathogenic in UKB-EUR (6.3%) compared to G&H (4.2%) (**Supplementary Table 5**). Accordingly, we found higher average number of pathogenic variants per gene (5.32 versus 3.6) and per individual (2.38 versus 2.19) in UKB-EUR compared to G&H (**Supplementary Figure 4B**). We found that the fraction of variants with unknown (VUS) or conflicting interpretations (CI) was also greater in UKB-EUR (56.4%) compared to G&H (39.1%) (**Supplementary Figure 4C**), in line with the observation among the South Asian samples in the RGC-ME dataset^11^. Interestingly, the portion of the benign variants was considerably lower in UKB-EUR (37.0%) compared to G&H (56.4%). It is possible that this difference reflects the differing practice of reporting or classifying variants by clinical sequencing labs in different countries. It is well recognized that variants in rare disease patients of non-European ancestry are harder to interpret due to the dearth of reference data^14,15^.

##### Homozygous genotypes in autosomal recessive disease genes

Among the 3,054 Mendelian disease genes with reported autosomal recessive (AR) inheritance, we found 368 genes (12%) with homozygous pLoF-HC genotypes and 2,060 genes (67%) with homozygous pDM genotypes in G&H (**Supplementary Figure 4D,E, top**). A large portion of the pLoF-HC and pDM variants with homozygous genotypes are absent in ClinVar (63% and 56%, respectively) or VUS/CI in ClinVar (12% and 29%, respectively) (**Supplementary Figure 4D,E, middle**). Among the pLoF-HC and pDM variants that are absent or VUS/CI in ClinVar, the majority (73% and 63%, respectively) did not have homozygous genotypes in UK Biobank or gnomAD (**Supplementary Figure 4D,E, bottom**), suggesting a unique opportunity that G&H provides to assess the clinical impact of these variants.

#### Supplementary Note 3: Genomic inflation and example of associations from rare variant association analyses

##### Genomic inflation

None of the single variant tests of the quantitative traits had inflation factors >1.1, and only phenotype ‘height’ had slightly inflated values with 1.13 and 1.12 for some of the gene-based masks, all other quantitative gene-based tests with AF<0.01 and excluding MASK A had inflation factors <1.12. Of the binary gene-based tests, there were some pregnancy traits and the trait ‘M75-Shoulder lesions’ with slightly elevated inflation factors <1.15 for AF<0.01 and excluding MASK A. For the binary single-variant tests there were 34 traits out of 681 traits with elevated inflation factors, the largest 1.32. However, all corresponding QQ plots looked normal and as expected.

##### Representative known associations

Quantitative associations: Gene-based associations between 13 genes (*ABCA1*, *ABCG5*, *ANGPTL3*, *APOA1*, *APOB*, *APOC3*, *CD36*, *CETP*, *LCAT*, *LDLR*, *LIPG*, *LPL*, *PCSK9*) and cholesterol levels; between 3 genes (*ANGPTL8, APOB, APOC3)* and triglyceride levels; between *GPLD1* and alkaline phosphatase levels; and between *GPT* and alanine aminotransferase.

Binary associations: We identified known associations such as *NF1* and benign neoplasms (ICD10 D36), *LDLR* and coronary artery disease/myocardial infarction, *SLC30A8* and type 2 diabetes, and *HNF4A* and type 2 diabetes^16-18^.

##### *APOB* and lipid traits

We identified one splice acceptor pLoF-HC (c.11904-2A>C, MAF=2.50x10^-4^, AC=22) with only one observed heterozygote in gnomAD associated with three cholesterol phenotypes and three distinct pDM’s absent from gnomAD NFE (p.Leu838Pro, MAF=1.14x10^-4^, AC=10; p.Ser3342Phe, MAF=1.10x10^-3^, AC=97) or >10x enriched in G&H (p.Arg532Trp, MAF=7.29x10^-3^, AC=642) associated with triglycerides or cholesterol traits.

##### *ABCB6* and serum potassium

We identified novel associations between three rare (MAF<0.2%) pDM variants in *ABCB6* and increased serum potassium levels. *ABCB6* encodes an erythrocyte membrane ABC transporter, and rare *ABCB6* missense variants have been reported to cause autosomal dominant pseudohyperkalaemia, a temperature-dependent passive leak of red blood cell potassium into plasma^19^. One of the variants associated with potassium levels was also associated with increased mean cell volume, consistent with a prior report of macrocytosis in an individual with pseudohyperkalaemia^19^, and may be a consequence of cellular swelling due to a disrupted equilibrium between intracellular and extracellular sodium and potassium^20,21^. One of the associated variants, p.Arg375Trp (chr2_219216028_G_A), is observed with MAF 3.4x10^-4^ in G&H (all heterozygotes) compared to 8.5x10^-7^ in gnomAD NFE. One previously reported pedigree for pseudohyperkalaemia was identified in East London ^22^ and correspondence with the authors suggest this is indeed the same extended family.

##### *FOLH1* and folate

We identified an association between the missense variant His475Tyr in the folate hydrolase *FOLH1* and serum folate levels. Previous studies have reported associations between variants in or near *FOLH1* and concentration of its alternative substrate N-acetyl-aspartyl-glutamate^23-25^, but this is the first report of an association with folate levels.

##### *TCN2* and vitamin B12

We identified associations between two predicted deleterious missense variants, Leu376Ser and Arg215Trp, in the vitamin B12 transporter *TCN2* and vitamin B12 levels. The missense variant Leu376Ser has been previously reported as associated with B12 levels, but the Arg215Trp association is novel. Notably, Arg215Trp is enriched in Genes and Health (MAF=2%) relative to non-Finnish Europeans in gnomAD (MAF=0.6%).

#### Supplementary Note 4: Sub-threshold associations from recessive burden analyses

We find a significant recessive association between *CLYBL* and vitamin B12 (p_rec_=1.26×10^-8^; beta=-0.48; p_domdev_=0.0069), recapitulating previous studies. 39 of 44 biallelic carriers in our cohort were homozygous for a pLoF variant in this gene (rs41281112) which has been already associated with deficiency of B group vitamins in UK Biobank^26^ (p_rec_=1.23×10^-12^). Importantly, Shen et. al^27^ demonstrated that CLYBL knockouts lead to deficiency of B12, in accordance with the large negative effect we detect. Furthermore, we highlight a few associations with binary phenotypes that, while not meeting the multiple-testing threshold, they show p_rec_<5.0×10^-6^ and p<0.05 for dominance deviation; a more detailed list of all associations with p_rec_<5.0×10^-6^ is given in **Supplementary Table 14**.

We observe a recessive association between *SKIV2L* and bacterial pneumonia (p_rec_=2.7×10^-6^; p_add_=0.0013; p_domdev_=0.0257), a gene located in the major histocompatibility complex with a direct role in immune response regulation^28^. Individuals homozygous or compound-heterozygous for pathogenic variants on *SKIV2L* exhibit Tricho-hepato-enteric syndrome, a rare autosomal recessive disease with complete penetrance, which is often associated with immune deficiency^29^, thus potentially explaining the observed association.

We associate *UGT2B15* with calculus of kidney and ureter (p_rec_=1.1×10^-6^; p_add_=7.4×10^-4^; p_domdev_=2.0×10^-4^). This gene is highly pleiotropic and encodes an enzyme involved in glucuronidation, a key process in the metabolism of various substances. Thus, individuals with biallelic pLoFs might have increased risk of kidney stones due to hyperoxaluria^30^. The above associations appear to be novel, as we found no direct support from previous GWASs, and would require replication in larger cohorts especially since they did not pass our formal significance threshold.

#### Supplementary Note 5: Implication of fine-scale population structure and relatedness for LoF-driven discoveries

The fine-scale population structure in G&H British Pakistanis has implications for the distribution of biologically interesting genotypes such as homozygous LoFs. Among the unrelated G&H Pakistanis, a pair of individuals with the same rare pLoF genotypes (AF<0.1%) have a ~27% chance of coming from the same cluster compared to only ~9% from randomly selected pairs (**Supplementary Figure 11A**). In Pakistani and Bangladeshi sub-samples with matched autozygosity, Pakistanis displayed a slightly lower rate of pLoF variants and genes with at least one homozygous pLoF genotype compared to Bangladeshis (p=4.3x10^-8^ and p<2.2x10^-16^, respectively) (**Supplementary Figure 11B**). Conversely, Pakistanis were more likely to carry pLoF variants for which there was more than one homozygous genotype (p<2.2x10^-16^) (**Supplementary Figure 11C**). This likely reflects slightly reduced genetic diversity, i.e. fewer pLoF variants, among the Pakistanis due to the bottlenecks in multiple subgroups and increased chances of the pLoF variants drifting to higher frequency due to these bottlenecks. This implies that if one aims to maximize the number of pLoF variants and genes with homozygous genotypes, sequencing more Bangladeshis would be more fruitful (conditional on a given level of autozygosity), whereas if the aim is to maximize the number of homozygous genotypes of certain pLoF variants, sequencing more Pakistanis would be more efficient.

We observed that individuals sharing the same rare homozygous pLoF genotypes (AF<0.1%) have ~30% chance of being first- or second-degree relatives, compared to <1% chance for randomly selected pairs (**Supplementary Figure 11D**). Thus, recall studies seeking to characterise the impact of knockouts could boost power by recruiting close relatives of index individuals. However, in the absence of a strong biological hypothesis, it could be difficult to definitively attribute a given phenotype to the homozygous pLoF in question rather than other shared genetic variation if one had only a small number of e.g. siblings with that genotype.

#### Supplementary Note 6: Characterization of genes with pLoF variant carriers

We evaluated what characteristics of the gene influence the likelihood of identifying heterozygous and biallelic pLoF genotypes. Adjusting for inherent features of genes such as CDS length, exon count, and tissue expression (**Supplementary Methods**), we found that genes that are essential in cell culture or knockout lethal in mice were depleted of both heterozygous and homozygous pLoF genotypes, with stronger depletion observed for the latter (**Supplementary Figure 10; Supplementary Table 16**). Genes that are haploinsufficient were depleted of heterozygous and homozygous pLoF genotypes to a similar degree, suggesting no additional effect of biallelic loss beyond monoallelic loss. While heterozygous and homozygous genotypes were similarly depleted among the autosomal dominant and monoallelic developmental disease genes, there was a contrasting pattern among the autosomal recessive and biallelic developmental disease genes: homozygous pLoF genotypes were depleted as expected while heterozygous pLoF genotypes were in fact enriched. This suggests that there may be a distinct evolutionary pressure on the occurrence of heterozygous versus homozygous loss of gene depending on the level of loss required for the gene to influence fitness.

### Supplementary Methods

#### Whole exome sequencing and variant and genotype calling

##### Sequencing and variant calling

The Broad Institute performed whole exome sequencing (WES) using Twist exome capture reagents on Illumina’s NovaSeq 6000 sequencer (150bp paired-end reads) following the 'Standard Germline Exome v6' protocol and the 34.9Mb Twist Alliance Clinical Research Exome (<https://www.twistbioscience.com/resources/safety-data-sheet/twist-alliance-clinical-research-exome-349-mb-bed-files>). BWA-MEM was used to map the reads to the reference genome (hg38) with ALT contigs to produce gVCF and CRAM files per sample. Preprocessing and variant calling were performed using the ExomeGermlineSingleSample 3.0.0 pipeline using Picard 2.23.8, GATK 4.2.2.0 HaplotypeCaller, and Samtools 1.11 (<https://broadinstitute.github.io/warp/docs/Pipelines/Exome_Germline_Single_Sample_Pipeline/README>). We obtained 48,737 CRAMs meeting >85% bases at >20x Twist bait target coverage. Variants on chromosome X were called assuming diploid for females and males while variants on chromosome Y and mitochondrial DNA were omitted. Note that chromosome X was not analysed in this paper.

##### Initial sample QC and joint calling

The following sample filters were applied sequentially in the following steps: 29 CRAMs failed due to having <85% bases at >20X coverage in GENCODE exons; 573 CRAMs failed the contamination estimate freemix >0.03; 55 CRAMs failed due to having self-stated gender for the individual that did not match the biological sex inferred from exome data (and could not be reconciled); 367 CRAMs were failed due to the individual having no valid NHS number (these also included study withdrawal, and incomplete consent); 3,414 CRAMs which were from an individual (based on unique NHS number) already sequenced (the lowest coverage CRAM(s) were removed). After this initial sample QC, 44,302 qualifying CRAMs proceeded to joint genotype calling using HAIL and GATK GenotypeGVCFs using the Broad Institute’s Joint Genotyping pipeline (<https://broadinstitute.github.io/warp/docs/Pipelines/JointGenotyping_Pipeline/README>)

##### Sample QC applied to VCF files

###### Comparison to chip genotype data

44,302 WES genotype data were compared to 44,396 Illumina GSAv3 chip (GSA-chip) genotype data^31^ to evaluate concordance among the 3,596 common (MAF>0.001) variants that were captured in both. There was a perfect or near-perfect match for 38,615 WES to GSA-chip pairs; 917 WES to GSA-chip pairs with high concordance had different Oragene IDs between WES and GSA-chip data (this means that the same person with a given NHS number has taken part twice or more with different Oragene IDs) and these were retained. We identified 9 pairs of identical twins, all of whom were retained in the WES dataset. In total, 37 WES samples with unresolvable discrepancy against the GSA-chip data were removed as likely recruitment or laboratory errors. Further sample QC was applied to the remaining 44,265 samples after variant calling, as described below.

###### Sex imputation

For sex imputation, the data were filtered to only include biallelic variants with MAF>0.05 and call rate>0.99. The sex of each sample was imputed using Hail’s impute_sex function with parameters male_threshold=0.79 and female_threshold=0.55, which calculates the inbreeding coefficient on the X chromosome. Comparison of the imputed sex to self-reported gender found discrepancies in eight samples which were flagged but not removed. Seven of these samples were previously known or suspected to have Klinefelter’s syndrome. The remaining sample, whose self-reported gender did not match the imputed sex based on Hail impute_sex function but matched the sex inferred based on cram Y total reads/X total reads on CRAM, was kept. 1,499 samples were identified as outliers, with an inbreeding coefficient (on X chromosome) of >0.2 and <0.8. These were almost all female samples with high autozygosity, suggesting that large autozygous regions on X chromosome likely interfered with sex imputation using the Hail impute_sex function. These samples were also flagged but not removed.

###### Inference of genetic ancestry for QC purposes

Variants were filtered to only include biallelic autosomal variants with MAF>0.001 and call rate>0.99. Variants in linkage disequilibrium were pruned using Hail’s ld_prune function with an r^2^ threshold of 0.2. The G&H samples were merged with reference samples from the 1,000 Genomes Project, and the variants present in both datasets were retained. Further filtering was performed by removing variants with low call rate (<0.99), low allele frequency (MAF<0.05), low Hardy-Weinberg equilibrium p-value (<1x10^-5^), variants in long-range linkage disequilibrium regions, and palindromic variants. Principal components (PC) were derived using Hail’s hwe_normalized_pca function. Superpopulation, i.e., continental-level population, of G&H individuals were predicted using gnomAD’s assign_population_pcs function. 106 samples were predicted not to be of South Asian ancestry and were excluded from the dataset.

After filtering the merged dataset to only include South Asian samples, UMAP was applied using the first 7 PCs (implemented in the python package umap-learn). Most of the samples were assigned to two clusters corresponding to the self-reported ancestries of Pakistani (17,793 samples) and Bangladeshi (26,169 samples). Similarity to the relevant 1000 Genomes Project populations also confirmed their ancestral assignment. Two minor clusters consisting of 198 samples were also retained.

###### Trio identification

To identify trios, GSA-chip data of 44,190 individuals were used^31^. We only included autosomal (chromosomes 1-22) and common (MAF >0.01) variants with call rate >99% and variants that passed HWE in declared Bangladeshi individuals (p<1x10^-6^). Trios were inferred with KING (version 2.3.0) up to 3rd degree relationships and filtered by allowing only plausible ages of parents and offspring, as well as low Mendelian error rates, i.e. families with >40 Mendelian errors per family were removed.

###### Sample QC based on variant metrics

Hail’s sample_qc function was applied, and the output stratified by the three ancestral groups: Bangladeshi, Pakistani, and other South Asians (as defined in the PCA analysis above). The following metrics were calculated per sample:

- Number of SNVs
- Ti/Tv ratio (transition/transversion ratio)
- Het/hom ratio (heterozygote/homozygote ratio)
- Heterozygosity rate
- Number of transitions
- Number of transversions
- Number of insertions
- Number of deletions
- Insertion/deletion ratio

Samples were removed if they had values outside the median ±6 median absolute deviations (MAD) within the given ancestry group for any metrics, except for het/hom ratio and heterozygosity rate, for which samples with values higher than the median +6 MADs were removed (to avoid removing samples with high autozygosity who had a low het/hom ratio). Overall, 44,028 individuals (24,444 females and 19,584 males) passed the population based (106 samples were removed) and variant metrics based (131 samples were removed) sample QC.

##### Variant QC applied to VCF files

A random forest model was trained with the aim to remove variants that are likely to be sequencing artefacts or mapping problems, while retaining as many true variants as possible. To minimise the number of variants with low coverage in off-target regions, only variants within the Twist bait regions (+/- 50bp) were used.

###### Preparation of truth set and false positives

The following variants were used as true positive variants.

- High confidence variant sites discovered in the 1,000 Genomes Project
- SNVs present on the Illumina Omni 2.5 genotyping array and found in the 1,000 Genomes Project
- INDELs present in the Mills and Devine data^32^
- SNVs and INDELs in HapMap3
- Variants failing a set of hard filters and deemed false positive: QD<2 or FS>60 or MQ<30

###### Train and apply random forest

A random forest model was trained on chromosome 20 using the true positive and false positive variants as described above and then applied to the whole dataset. The feature choice was largely based on the set of features used by the gnomAD project. Here additionally, meanHetAB (mean allele balance, i.e., ALT/REF reads, at heterozygous genotypes) was used to identify and subsequently remove potential artefacts (**Supplementary Table 20**). Note that some variants were annotated as FILTER=ExcessHet at the alignment stage by the Broad Institute. These variants were not removed if they passed the random forest QC.

The random forest model assigns a score to each variant, i.e., the likelihood of being a true variant. To determine a truth threshold which impacts the sensitivity and specificity of the final variant callset, variants were first ranked by their random forest score and assigned to bins. Then plots of cumulative true positive variants per bin versus cumulative false positive variants per bin were inspected for both SNVs and INDELs, as well as plots of transmitted/untransmitted ratio for synonymous singletons (SNVs only) in combination with various genotype quality metrics (described below). The transmitted/untransmitted ratio for synonymous singletons makes use of the trios (see below) in the dataset (N=471) and examines synonymous variants seen in only one parent in the dataset. If the QC is properly calibrated, such variants are expected to be transmitted to the child 50% of the time. Hence, the QC thresholds should be chosen so that this metric is close to 1 while also optimising the other metrics outlined in the next section.

##### Genotype QC

###### Autosomes

Different combinations of random forest bins (i.e., variant-level metrics) were analysed together with various genotype quality metrics such as DP (depth), GQ (genotype quality) and HetAB (heterozygous allele balance, i.e., the fraction of reads carrying the alternate allele at a heterozygous genotype). Specifically, if variants passed a given random forest bin filter, genotypes were set to missing if they did not pass one or more of the GQ, DP or HetAB thresholds. For each filter combination various metrics were calculated, i.e. the percentage of true positive and false positive variants, the transmitted/untransmitted synonymous singleton ratio in the trios, the total number of Mendelian errors in the trios, and the mean number of heterozygous calls in ROHs (runs of homozygosity) called using bcftools roh from the GSA genotype chip data. Additionally, several call rate filters (call rate >0%, >50%, >90%, >95%) were evaluated at the same time.

###### Chromosome X

Although both sexes were called using a diploid model on the X chromosome (since this was the default for the Broad variant-calling pipeline), males are hemizygous in the non-PAR regions of chromosome X so we tested different genotype-wise DP and GQ thresholds separately for males and females. After applying a combination of filters, the same metrics as described previously were calculated, apart from the number of heterozygous calls in ROHs, which was replaced by the mean number of heterozygous calls in the male’s non-PAR regions.

###### Final choice of variant and genotype filters

Three sets of filters were tested at different levels of strictness and their performance evaluated (**Supplementary Table 21** for SNVs and **Supplementary Table 22** for INDELs). Having considered the various options and performance, we decided to use the stringent filter for downstream analyses. Specifically, for SNVs, we removed variants in random forest bin > 80, or with <95% genotypes non-missing after applying the following filters: DP <10, GQ<20, AB<0.2. For INDELs, we removed variants in random forest bin > 44, or with <95% genotypes non-missing after applying the following filters: DP<10, GQ<20, AB<0.3, For the non-PAR region on chromosome X in males, the DP filter was relaxed to 5 for SNVs, but it was kept at 10 for INDELs. Metrics obtained using the stringent filters are shown in **Supplementary Table 23**.

##### Phasing

We performed statistical phasing in two steps, following approaches described in the recent studies of UK Biobank^33-35^. We merged the exome and array genotypes, resulting in a set of ~4.7 million variants and 39,320 individuals. Using SHAPEIT5^34^, we first phased common variants (MAF>0.001), then used the resulting haplotypes as a scaffold to phase rare variants, in chunks of 4cM length. SHAPEIT5 is efficient for large samples and suitable for rare variants, as it provides a confidence score for each genotype termed phasing probability (PP; note that PP=1.0 when MAF>0.001). We used the PP values to assess our phasing approach and to select genotype for downstream analyses.

A standard metric used to assess phasing accuracy is switch error rate (SER) calculated in parents-offspring trios by comparing statistically phased genotypes to genotypes inferred by Mendelian inheritance in the offspring. We examined SER in 100 trios (100 offspring and 172 parents; some parents have >1 offspring) to assess phasing accuracy. In the remaining subset (n=39,148) that was used for downstream analyses, we surveyed all rare variants that were transmitted from one parent and phased and found SER of 0.55% (**Supplementary Figure 8A**). Reassuringly, SER dropped to 0.34% among the genotypes with high-confidence phasing (PP>0.9) which were used for downstream analyses.

#### Extraction and preparation of phenotypes from electronic health record data

##### Quantitative traits

Quantitative traits were extracted from all available electronic healthcare record (EHR) data sources, including data from London based primary care, Barts Health NHS Trust, and Bradford Teaching Hospitals NHS Trust. Because several quantitative traits are strongly influenced by acute illnesses, we elected to use data from the primary care sources only, excluding secondary (hospital) care data. We extracted a broad range of quantitative traits from the primary care records in up to ~54,000 volunteers of the Genes & Health Study. We applied a stringent stepwise quality control procedure to derive individual-level phenotype data from the EHR. Briefly, after harmonising EHR data across the different data sources, we excluded test results which were non-numeric. Test results which contained ">” or "<” indicators were simplified to represent this limit, e.g., a C-reactive protein of <1 was relabeled as 1 (note that we subsequently only use non-parametric statistics). We then used custom code lists to define all occurrences of a test result in the EHR. For each trait, we defined a desired target unit and converted all test results for the trait to match the target unit. Test results with incompatible units, i.e., units which could not be converted with a simple multiplication factor, were excluded. We defined a manually curated minimum and maximum plausible value range for each trait influenced by prior clinical knowledge and reported ranges of these traits in UK Biobank (where available). It is important to note that these ranges were devised to exclude values which are probably due to technical errors in sample processing or data entry, whilst retaining as much biological variation as possible. They are therefore somewhat deliberately broad. Trait values above or below the manually specified a priori limits of plausibility were excluded. Test results obtained from before the age of 18, labelled as after the date of data extraction, or with a missing date were excluded. The age at test was approximated from the month and year of birth stated by the volunteer in their baseline questionnaire. It is possible that a single test result appears in the EHR multiple times due to duplicate data entry mechanisms. For this reason, we excluded test results with the exact identical value occurring within a 10-day rolling window.

To account for the impact of medications on quantitative traits, we first cleaned and curated prescription data from the primary care EHR. We then manually defined a set of commonly prescribed medications which can influence the traits studied in this paper and defined the earliest prescription date per-individual per-drug. Rather than adjusting the values obtained while 'on-drug', we chose the pragmatic approach of restricting to only data points obtained prior to drug initiation. The following drugs were accounted for: anti-hyperglycaemic agents (insulins, metformin, gliclazide, GLP-1 agonists, SGLT2 inhibitors) for glycaemic traits (HbA1c, random glucose), B12 (for vitamin B12), vitamin D (for serum vitamin D), folate (for serum folate), iron (for serum ferritin), statins (for cholesterol, LDL-C, HDL-C, and triglycerides), and thyroxine (for T4 and TSH).

Blood test results in hospitalised inpatients are more likely to reflect transient changes related to acute illnesses and so are less likely to be a reflection of steady-state biology. While the overall population mean values for primary and secondary care traits were highly correlated, there was substantially greater heterogeneity in the secondary care data. We therefore restricted the dataset to readings from primary care data. Following exclusion of secondary care readings, we excluded outlying values more than 10-standard deviations from the mean on the log-10 scale. This standard deviation threshold was chosen through an iterative process which aimed to maximise specificity (i.e., minimise the risk of including dubious test results reflecting errors in data entry, units, or failed assays) while retaining true biological extremes. The resulting trait distributions were largely normally distributed on the log-10 scale, with a small number of exceptions (eosinophils, CRP, and ESR). Intra-individual variation was low consistent with recent descriptions of set-points for blood traits^36^.

As both age at test and year of test explained a non-trivial amount of variation in several traits studied, we used a regression-based approach to account for these covariates upstream of exome-wide association testing. We first identified the median reading per individual and used this reading to obviate issues with inter-dependence of multiple readings from the same person. We used linear regression models adjusted for age at test, age at test^2^, year of test, and year of test^2^ to account for the effects of age at test and year of test. While the impact of age is likely to reflect both biological (i.e., age-related changes) and confounding effects, the impact of year of test is more likely to reflect confounding effects, e.g., changes in laboratory assays over time. Importantly, in many cases the age at test differed substantially from the age at recruitment owing to the longitudinal nature of the healthcare record linkage. To ensure that model assumptions were satisfied, the outcome for the models was the log10-transformed trait value standardized by z-scoring. Applying this model to the post-quality control individual-level data yielded a residual trait value for each individual (corresponding to that individual's lifetime median, adjusted for age at test and year of test). Visual inspection of residual plots for each trait confirmed that this approach preserved the overall structure of the data. The median residual per person was then further transformed using rank-inverse normalisation prior to association testing. Importantly, this procedure preserved the ranking of the distribution while destroying the underlying distribution on the original scale, and so the beta coefficients from association studies cannot be interpreted as an absolute effect size.

##### Binary phenotypes

Binary traits were defined using data from all available electronic healthcare record (EHR) data sources, including data from London based primary care, Barts Health NHS Trust, Bradford Teaching Hospitals NHS Trust, and NHS England (Cancer, Mortality, Hospital Episode Statistics). Multiple (approximately annual) refreshes of data were merged and de-duplicated.

We used TREtools (<https://github.com/genes-and-health/tre-tools>) for our data processing and binary phenotype generation. TREtools is an open-source Python package that simplifies the process of running code lists against various datasets, enabling users to quickly identify relevant patient data using SNOMED, ICD10, and OPCS code lists. The package can also convert between standard code list formats (e.g., from SNOMED to ICD10) using a linkage file. Additionally, it offers thorough data cleaning and processing functions, transforming raw data into ready-to-use formats by standardising columns, removing duplicates, and aligning date formats. The package includes various test modules to maintain data quality and accuracy, validating code lists and checking individual counts during processing, which helps ensure data integrity throughout the analysis. The package also supports creating phenotype reports to analyse patient demographics, calculate specific conditions, and identify overlaps across datasets, making it a valuable tool for efficient and structured healthcare data analysis.

We generated ‘first occurrence of 3-digit ICD10’ binary traits, using methods as similar as possible to what was previously done by UK Biobank (<https://biobank.ndph.ox.ac.uk/ukb/ukb/docs/first_occurrences_outcomes.pdf>). NHS England Cancer Registry data was used for G&H, while it was not in the UK Biobank. ICD10 codes (3- or 4-digit depending on dataset) were obtained from secondary (hospital) care data sources. SNOMED codes were also obtained from some hospital datasets and primary care and were mapped to 4-digit ICD10 (and then reduced to 3-digit ICD10) codes using mapping files from NHS England (der2_iisssccRefset_ExtendedMapFull_INT_20210131.txt; der2_iisssciRefset_ExtendedMapUKCLFull_GB1000000_20220413.txt). SNOMED codes that mapped to multiple 4-digit ICD10 codes were discarded (otherwise we found spuriously large numbers of cases of anthrax and various other infectious diseases as also noted by UK Biobank). Multiple SNOMED codes mapping to a single 4-digit ICD10 code were kept. 4-digit ICD10 codes were then reduced to 3-digit, and date, age, and dataset were recorded for the earliest occurrence of the code for each individual. 1,833 3-digit ICD10 coded traits (beginning A to Q – codes after Q are administrative not disease codes) were generated. For REGENIE association analyses, 3-digit ICD10 coded traits with <100 individuals were omitted. The 3-digit ICD10 traits are intended as a broad sweep across all possible human phenotypes. Additionally, we compiled 248 custom binary traits based on 3-digit ICD10, 4-digit ICD10, SNOMED and OPCS code lists compiled by multiple researchers. OPCS coded data were not used in the 3-digit ICD10 phenotypes. An exact full word match to the code in the code list to the code in the health dataset was required for inclusion. Date, age, and dataset were recorded for the earliest occurrence of the code for each individual. Coded traits with <100 individuals were omitted from association analyses. The full code lists can be found in **Supplementary Table 25**.

#### Fine-scale population structure and founder variants

##### Inference of genetic ancestry

Note that this section describes the inference of genetic ancestry for the plots shown in Supplementary Figure 1, which then fed into the fine-scale population structure analyses described in the next section. It is distinct from the ancestry inference done using Hail which was done for the purposes of sample and variant QC, as described above. We repeated this broad-scale ancestry inference using the GSA data rather than the pre-QC exome data since we wanted to bring in reference samples from other datasets for which whole-genome sequence data were available, and genome-wide common variants were likely to improve our classification of ancestry.

To identify Bangladeshi and Pakistani ethnicity, we used the GSA-chip genetic data^31^ of 51,176 Genes & Health individuals and 637,829 variants. Of those 51,176 individuals, 43,507 have both WES and GSA data. To merge the GSA-51k call set with a reference genome set, alleles were flipped to the positive strand by running the script HRC-1000G-check-bim-NoReadKey.pl (Will Rayner, <https://www.strand.org.uk/>) which resulted in 507,357 variants. After the following filtering steps 347,606 variants remained: only autosomal (chromosomes 1-22) and common variants (MAF >0.01) were included with call rate >99% and HWE p-value <10^-6^ in declared 26,174 Bangladeshi individuals. The reference genome set consists of 3,433 individuals from the 1000 Genomes Project^37-39^ (1000GP Phase 3; 2,504 individuals; <https://www.internationalgenome.org/1000-genomes-summary>) and the Human Genome Diversity Project^40^ (HGDP; 929 individuals) which shared 346,513 variants with the Genes & Health cohort.

Principal components analysis (PCA) was performed on 103,438 variants after LD pruning (window size 1000kb, step size 50, and LD r2 cutoff 0.1) and excluding long LD regions (<https://github.com/meyer-lab-cshl/plinkQC/blob/master/inst/extdata/high-LD-regions-hg38-GRCh38.txt>).

PCA was used in two steps to infer ethnicity. In the first round, the first twenty principal components (PCs) were used to project the 51,176 Genes & Health individuals onto the reference PCA space (Figure S1). Uniform Manifold Approximation and Projection for Dimension Reduction (UMAP) using eight PCs showed a clean separation of the reference super populations and the highest combined sensitivity and specificity for inferring Bangladeshi and Pakistani ethnicities. Therefore, UMAP with eight PCs were used to infer Bangladeshi and Pakistani individuals, and six outliers were identified and excluded (labelled ‘Others’). After excluding the six population outliers, 35,146 unrelated individuals up to 2nd degree (KING v2.3.2) were identified, and in the second round, PCA was repeated on these unrelated individuals (**Supplementary Figure 2**). The remaining 16,024 related individuals were projected onto the PC space of unrelated individuals, using the same 103,438 variants. UMAP with six PCs identified distinct Pakistani and Bangladeshi clusters and a third small cluster without teasing out excessive population structure. 64 individuals (labelled ‘Ambiguous’) switched inferred ethnicities from the first PCA/UMAP mapping using the reference datasets, but they did not form their own cluster.

##### Deriving fine-scale population structure

To investigate fine-scale population structure, principle component analysis (PCA) was carried out for 51,176 Genes & Health (G&H) individuals with Illumina Global Screening Array (GSA) data together with a South Asian reference panel of 1,864 individuals from the ‘Born in Bradford’ study (BiB), 187 Pakistanis from HGDP and 489 South-Asians (SAS) from 1000GP (Phase 3), resulting in 53,716 individuals in total (**Supplementary Figure 1**). The same South Asian reference panel was also used further to identify population fine-structure in the Pakistani population using identity-by-descent-based clustering, as we now describe. As a first step, KING (v2.3.2) was run for the whole set of 53,716 individuals to identify a more stringent set of unrelated individuals. The options --unrelated and --degree 3 in KING generated a ‘segments’ file which was used to remove individuals with the most relatives until no relationship pair with degree of 3 or closer remained. If there was a tie in the number of relationships, individuals with WES data were kept preferentially. This procedure resulted in 32,152 unrelated individuals, of which 26,957 had WES data. Second, IBIS^41^ was run for the set of 53,716 individuals to obtain shared IBD segments >5cM (options -ibd2 -t 2 -mL 5 -mt 500 -er 0.004). Finally, if an unrelated pair as defined by KING shared an IBD segment >40cM, one of each pair was removed, keeping preferentially individuals with WES data, which resulted in 20,438 individuals (19,390 G&H, 461 BiB, 148 Pakistani from HGDP and 439 SAS from 1000GP) and 17,172 G&H individuals with WES data. Of those unrelated 17,172 G&H individuals, 8,109 were inferred Pakistani. We removed BiB individuals from subgroups in which fewer than five individuals remained after this relatedness filtering, leaving 121 individuals. Then a graph (igraph in R) was constructed with edge weights corresponding to the total length of IBD segments shared between each pair of individuals, and the Louvain method (a hierarchical clustering algorithm) was applied to detect community structure using these shared IBD segments (resolution 1.4; igraph package in R). The reference populations were clustered together with the G&H Pakistanis, which generated 23 clusters. Two of the clusters contained fewer than five G&H Pakistanis and were removed. IBD scores were calculated as the total length of IBD segments shared between any two individuals within a cluster, normalised by the pairs of individuals within a cluster^42^.

##### Founder effects

To establish whether there are any founder effects in Pakistani sub-populations, i.e. whether there are variants with allele frequencies (AF) significantly elevated in any of the clusters, Fisher’s exact tests were carried out for each cluster (=cases) versus all other clusters (=controls). To determine the AF at which we had power to identify enrichment and to determine a p-value significance threshold, allele counts (AC) for all cases and controls combined were varied, starting with AC=1, AC=2, etc. Then for each given AC, Fisher’s exact tests were applied for different odds-ratios, i.e. AC for the ‘case’ cluster were permuted from AC=1 to the maximum AC of cases and controls combined. For each overall AC, the number of association tests was calculated, and Bonferroni p-value thresholds were computed by dividing the nominal p-value by the total number of tests carried out across all clusters. For each cluster, the minimum AC was identified for which any Bonferroni corrected p-value was lower than the Fisher’s exact test p-values. Association tests with AC lower than the minimum AC were discarded since the power was deemed too low to detect meaningful associations. After determining the minimum AC for each cluster, an overall Bonferroni corrected p-value significance threshold was calculated as p <5.25x10^-9^. Cluster sizes and minimum AC can be found in **Supplementary Table 1**. We found that the number of founder variants can be modelled by fitting a linear model to the log-transformed counts of founder variants versus IBD scores and log-transformed sample size within a cluster as explanatory variables (**Supplementary Table 3**). An interaction term between IBD scores and log-transformed sample sizes was also significant, implying that for higher IBD scores there is a positive correlation between sample size and number of founder variants. This positive correlation disappears for very low IBD scores.

##### Comparison of homozygous pLoF genotype distributions between Pakistani and Bangladeshi individuals

To inform future study designs that aim to analyze loss-of-function variants (LoF), we sought to establish differences in homozygous pLoF genotype distributions between Bangladeshis and Pakistanis. Noting that Pakistanis have higher autozygosity than Bangladeshis^43^, we first sampled Bangladeshi and Pakistani individuals to match the level of autozygosity using the fraction of runs of homozygosity (F_ROH_). F_ROH_ was calculated with PLINK using array genotype data (Illumina Global Screening array) and split into 50 bins. We sampled an equal number of unrelated Bangladeshis and Pakistanis from each bin, i.e., the maximal number available in each population for each bin. In total, we sampled 5,172 unrelated individuals from each population 1,000 times, each time recording their homozygous pLoF genotypes with AF<1%. From that we calculated the number of genes and variants with pLoF genotypes, and the number of carriers of each pLoF variant in each population.

#### Rare variant association analyses (RVAS)

Principal components (PCs) used as covariates were calculated for common (MAF>1%) and independent (PLINK --indep-pairwise 50 5 0.5) variants among unrelated individuals (KING --degree 2). Related individuals were projected onto the PC space of unrelated individuals.

##### Permutation approach to determine p-value thresholds

To determine p-value significance thresholds, we permuted the genotypes once for each phenotype and calculated false discovery rates (FDR) for various p-value thresholds similar to the strategy previously described^26^. Wang et al.^26^ permuted phenotypes among unrelated individuals for Fisher’s exact tests and linear regressions. When using linear regressions, the noise term is assumed to be uncorrelated between individuals and there is no relationship adjustment that would be destroyed. However, our analyses included related individuals as Genes & Health has substantial relatedness and retaining related individuals boosts sample size. Since we adjusted for relatedness in the first step of the REGENIE analyses, it is more appropriate to permute genotypes instead of phenotypes in the second step of the REGENIE analyses. Using an FDR of 5%, the significance thresholds for quantitative traits were p<7.5x10-9 for single variant tests and p<4.4x10^-7^ for gene-based tests. For binary traits, the significance thresholds were p<3.3x10^-8^ for single variant tests and p<3.5x10^-8^ for gene-based tests.

##### Conditional analyses using common variants from GWAS

To derive single variant and gene-based associations from RVAS that are independent of nearby common variant associations, we performed conditional analyses as follows. We used summary statistics from genome-wide association studies performed with imputed genotypes in G&H derived based on GSA-chip genotypes and TOPMed-r3 reference panel^31^. We identified common variants with nominal association (p<1x10^-6^) within 5Mb of the variants or genes with significant associations from RVAS. To arrive at an independent set of nominally significant common variants, we LD-pruned (r2<0.1) the GWAS variants using the LD estimated among unrelated (>3rd degree) participants in G&H. Finally, for each significant single variant and gene-based association which had nearby nominally significant common variants, we repeated REGENIE step 2 with identical parameters and covariates but with the addition of the dosage of the independent GWAS variants as additional covariates. Single variant or gene-based associations which retained p<0.05 after conditioning were considered independent.

##### Deriving novel associations

Variants were assigned to genes using the same annotations as the gene-based tests. We assigned variants that were not included in the gene-based tests to the gene(s) with the most severe consequence among MANE Select transcripts; 91 variants that did not have consequences for MANE Select transcripts were not assigned to genes. We mapped G&H phenotypes to EFO codes to facilitate comparison with public databases. Quantitative traits were manually mapped to EFO codes. Binary ICD10 first occurrence phenotypes were mapped to EFO codes using the EFO-UKB-mappings repository (https://github.com/EBISPOT/EFO-UKB-mappings) from EBISPOT. For ICD10 phenotypes without matches in EFO-UKB-mappings and phenotypes with custom code lists, phenotype names were searched in the Ontology Lookup Service and GWAS Catalog to identify appropriate EFO codes. 48 of 49 quantitative traits with significant associations and 118 of 127 binary phenotypes with significant associations were mapped to EFO codes (Supplementary Table 10).

To assess whether gene-phenotype pairs were likely “novel” or “reported” in previous GWAS/RVAS studies or Mendelian disease, we obtained published gene-based associations using the Ensembl ID and EFO ID from Open Targets Platform (version June 2024, 24.06). Locus2Gene (L2G) scores were obtained from Open Targets Genetics (version October 2022, 22.10) using Ensembl IDs and EFO IDs. We also assessed novelty by free text searching each gene-phenotype pairs Open Targets Platform (version June 2024, 24.06); Online Mendelian Inheritance in Man, OMIM (version October 2024, 24.10); and genebass, which reports gene-based associations and single-variant associations from the UK Biobank (version October 2024, 24.10). Associations from Open Targets Platform were filtered to variants with an L2G score > 0.5. A narrow definition and a broad definition of novelty were used. Under the narrow definition, gene-phenotype pairs were novel if the exact gene-phenotype pair association was not identified in any of the databases. Under the broad definition, associations with closely related phenotypes were also considered. For example, the gene *MAT2A* was associated with acute myocardial infarction in Genes & Health. This specific gene-pair association was found to be novel when searching databases, however previous associations between *MAT2A* and the closely related phenotype coronary artery disease were found. Thus, the gene-phenotype pair is novel under the narrow definition but not the broad definition. Gene-phenotype novelty and significance across the RVAS, meta-analysis, and recessive analyses are summarized in **Supplementary Table 10**.

#### Enrichment analyses for genes with pLOF carriers

Fisher’s exact test and logistic regression test were used to evaluate the relationship between gene features or gene sets and whether the gene has heterozygous or homozygous pLOF variant carriers in G&H. First, we examined the effect of the inherent features of the genes such as CDS length, exon count, gene expression - number of tissues expressed at TPM>=1 (quantitative) or expressed in all, some, or none of the tissues (categorical), protein localization – secreted, membrane, or intracellular (categorical), and gene family (categorical) on the presence of pLOF carriers (**Supplementary Table 16**). Significant gene features were then added as covariates in the downstream analyses. Next, we tested whether the genes with pLOF carriers were enriched or depleted in various functional or disease gene sets (**Supplementary Table 16**). CDS length and exon count are derived from Ensembl v105, gene expression from GTEx^44^ (v8), protein localization from HPA^45^, gene family information from IUPHAR/BPS^46^, essential genes from three cell culture screens^47-49^ through dbNSFP^50,51^ (v4.4c), mouse knockout lethal genes from IMPC^52^, haploinsufficient genes from ClinGen^53^, disease genes from OMIM^54^ and DDD^55^.

#### Code availability

Pipeline for phasing and identifying biallelic genotypes: <https://github.com/BRaVa-genetics/snakemake_pipeline_for_phasing>

Scripts for recessive association and other analyses: <https://github.com/giorkala/gnh_flagship>

TREtools for trait extraction and preparation: <https://github.com/genes-and-health/tre-tools>

### Supplementary References

1.	Cirulli, E.T. *et al.* Genome-wide rare variant analysis for thousands of phenotypes in over 70,000 exomes from two cohorts. *Nat Commun* **11**, 542 (2020).

2.	Piertney, S.B. & Oliver, M.K. The evolutionary ecology of the major histocompatibility complex. *Heredity (Edinb)* **96**, 7-21 (2006).

3.	Kwiatkowski, D.P. How malaria has affected the human genome and what human genetics can teach us about malaria. *Am J Hum Genet* **77**, 171-92 (2005).

4.	Wang, G.D. *et al.* Genetic convergence in the adaptation of dogs and humans to the high-altitude environment of the tibetan plateau. *Genome Biol Evol* **6**, 2122-8 (2014).

5.	Beall, C.M. *et al.* Natural selection on EPAS1 (HIF2alpha) associated with low hemoglobin concentration in Tibetan highlanders. *Proc Natl Acad Sci U S A* **107**, 11459-64 (2010).

6.	Simonson, T.S. *et al.* Genetic evidence for high-altitude adaptation in Tibet. *Science* **329**, 72-5 (2010).

7.	Yi, X. *et al.* Sequencing of 50 human exomes reveals adaptation to high altitude. *Science* **329**, 75-8 (2010).

8.	Arciero, E. *et al.* Demographic History and Genetic Adaptation in the Himalayan Region Inferred from Genome-Wide SNP Genotypes of 49 Populations. *Mol Biol Evol* **35**, 1916-1933 (2018).

9.	Landrum, M.J. *et al.* ClinVar: improvements to accessing data. *Nucleic Acids Res* **48**, D835-D844 (2020).

10.	Miller, D.T. *et al.* ACMG SF v3.2 list for reporting of secondary findings in clinical exome and genome sequencing: A policy statement of the American College of Medical Genetics and Genomics (ACMG). *Genet Med* **25**, 100866 (2023).

11.	Sun, K.Y. *et al.* A deep catalogue of protein-coding variation in 983,578 individuals. *Nature* **631**, 583-592 (2024).

12.	Venner, E. *et al.* The frequency of pathogenic variation in the All of Us cohort reveals ancestry-driven disparities. *Commun Biol* **7**, 174 (2024).

13.	Rony, R., Deng, S., Yang, S., Doig, K. & Goode, D.L. Putative breast cancer risk variants from populations of South Asian ancestry are under-represented in public variant classification databases. (medRxiv, 2025).

14.	Wright, C.F. *et al.* Genomic Diagnosis of Rare Pediatric Disease in the United Kingdom and Ireland. *N Engl J Med* **388**, 1559-1571 (2023).

15.	Tallman, S. *et al.* Missing genetic diversity impacts variant prioritisation for rare disorders. (medRxiv, 2024).

16.	Backman, J.D. *et al.* Exome sequencing and analysis of 454,787 UK Biobank participants. *Nature* **599**, 628-634 (2021).

17.	Karczewski, K.J. *et al.* Systematic single-variant and gene-based association testing of thousands of phenotypes in 394,841 UK Biobank exomes. *Cell Genom* **2**, 100168 (2022).

18.	Mahajan, A. *et al.* Refining the accuracy of validated target identification through coding variant fine-mapping in type 2 diabetes. *Nat Genet* **50**, 559-571 (2018).

19.	Andolfo, I. *et al.* Missense mutations in the ABCB6 transporter cause dominant familial pseudohyperkalemia. *Am J Hematol* **88**, 66-72 (2013).

20.	Haines, P.G. *et al.* Familial pseudohyperkalaemia Chiswick: a novel congenital thermotropic variant of K and Na transport across the human red cell membrane. *Br J Haematol* **112**, 469-74 (2001).

21.	Petrowsky, H. & Clavien, P.A. Chapter 44 - Principles of Liver Preservation. in *Transplantation of the Liver* 582-599 (ELSEVIER, 2015).

22.	Gore, D.M. *et al.* Four pedigrees of the cation-leaky hereditary stomatocytosis class presenting with pseudohyperkalaemia. Novel profile of temperature dependence of Na+-K+ leak in a xerocytic form. *Br J Haematol* **125**, 521-7 (2004).

23.	Feofanova, E.V. *et al.* A Genome-wide Association Study Discovers 46 Loci of the Human Metabolome in the Hispanic Community Health Study/Study of Latinos. *Am J Hum Genet* **107**, 849-863 (2020).

24.	Schlosser, P. *et al.* Genetic studies of urinary metabolites illuminate mechanisms of detoxification and excretion in humans. *Nat Genet* **52**, 167-176 (2020).

25.	Luo, S. *et al.* Genome-wide association study of serum metabolites in the African American Study of Kidney Disease and Hypertension. *Kidney Int* **100**, 430-439 (2021).

26.	Wang, Q. *et al.* Rare variant contribution to human disease in 281,104 UK Biobank exomes. *Nature* **597**, 527-532 (2021).

27.	Shen, H. *et al.* The Human Knockout Gene CLYBL Connects Itaconate to Vitamin B(12). *Cell* **171**, 771-782 e11 (2017).

28.	Eckard, S.C. *et al.* The SKIV2L RNA exosome limits activation of the RIG-I-like receptors. *Nat Immunol* **15**, 839-45 (2014).

29.	Fabre, A. *et al.* SKIV2L mutations cause syndromic diarrhea, or trichohepatoenteric syndrome. *Am J Hum Genet* **90**, 689-92 (2012).

30.	Dawson, C.H. & Tomson, C.R. Kidney stone disease: pathophysiology, investigation and medical treatment. *Clin Med (Lond)* **12**, 467-71 (2012).

31.	Jacobs, B.M. *et al.* Genetic architecture of routinely acquired blood tests in a British South Asian cohort. *Nat Commun* **15**, 8929 (2024).

32.	Mills, R.E. *et al.* An initial map of insertion and deletion (INDEL) variation in the human genome. *Genome Res* **16**, 1182-90 (2006).

33.	Barton, A.R., Sherman, M.A., Mukamel, R.E. & Loh, P.R. Whole-exome imputation within UK Biobank powers rare coding variant association and fine-mapping analyses. *Nat Genet* **53**, 1260-1269 (2021).

34.	Hofmeister, R.J., Ribeiro, D.M., Rubinacci, S. & Delaneau, O. Accurate rare variant phasing of whole-genome and whole-exome sequencing data in the UK Biobank. *Nat Genet* **55**, 1243-1249 (2023).

35.	Lassen, F.H. *et al.* Exome-wide evidence of compound heterozygous effects across common phenotypes in the UK Biobank. *Cell Genom* **4**, 100602 (2024).

36.	Foy, B.H. *et al.* Haematological setpoints are a stable and patient-specific deep phenotype. *Nature* **637**, 430-438 (2025).

37.	Genomes Project, C. *et al.* A map of human genome variation from population-scale sequencing. *Nature* **467**, 1061-73 (2010).

38.	Genomes Project, C. *et al.* An integrated map of genetic variation from 1,092 human genomes. *Nature* **491**, 56-65 (2012).

39.	Genomes Project, C. *et al.* A global reference for human genetic variation. *Nature* **526**, 68-74 (2015).

40.	Cavalli-Sforza, L.L., Wilson, A.C., Cantor, C.R., Cook-Deegan, R.M. & King, M.C. Call for a worldwide survey of human genetic diversity: a vanishing opportunity for the Human Genome Project. *Genomics* **11**, 490-1 (1991).

41.	Seidman, D.N. *et al.* Rapid, Phase-free Detection of Long Identity-by-Descent Segments Enables Effective Relationship Classification. *Am J Hum Genet* **106**, 453-466 (2020).

42.	Nakatsuka, N. *et al.* The promise of discovering population-specific disease-associated genes in South Asia. *Nat Genet* **49**, 1403-1407 (2017).

43.	Malawsky, D.S. *et al.* Influence of autozygosity on common disease risk across the phenotypic spectrum. *Cell* **186**, 4514-4527 e14 (2023).

44.	Consortium, G.T. The Genotype-Tissue Expression (GTEx) project. *Nat Genet* **45**, 580-5 (2013).

45.	Thul, P.J. *et al.* A subcellular map of the human proteome. *Science* **356**(2017).

46.	Harding, S.D. *et al.* The IUPHAR/BPS Guide to PHARMACOLOGY in 2024. *Nucleic Acids Res* **52**, D1438-D1449 (2024).

47.	Wang, T. *et al.* Identification and characterization of essential genes in the human genome. *Science* **350**, 1096-101 (2015).

48.	Hart, T. *et al.* High-Resolution CRISPR Screens Reveal Fitness Genes and Genotype-Specific Cancer Liabilities. *Cell* **163**, 1515-26 (2015).

49.	Blomen, V.A. *et al.* Gene essentiality and synthetic lethality in haploid human cells. *Science* **350**, 1092-6 (2015).

50.	Liu, X., Jian, X. & Boerwinkle, E. dbNSFP: a lightweight database of human nonsynonymous SNPs and their functional predictions. *Hum Mutat* **32**, 894-9 (2011).

51.	Liu, X., Li, C., Mou, C., Dong, Y. & Tu, Y. dbNSFP v4: a comprehensive database of transcript-specific functional predictions and annotations for human nonsynonymous and splice-site SNVs. *Genome Med* **12**, 103 (2020).

52.	Dickinson, M.E. *et al.* High-throughput discovery of novel developmental phenotypes. *Nature* **537**, 508-514 (2016).

53.	Rehm, H.L. *et al.* ClinGen--the Clinical Genome Resource. *N Engl J Med* **372**, 2235-42 (2015).

54.	Amberger, J.S., Bocchini, C.A., Schiettecatte, F., Scott, A.F. & Hamosh, A. OMIM.org: Online Mendelian Inheritance in Man (OMIM(R)), an online catalog of human genes and genetic disorders. *Nucleic Acids Res* **43**, D789-98 (2015).

55.	Deciphering Developmental Disorders, S. Large-scale discovery of novel genetic causes of developmental disorders. *Nature* **519**, 223-8 (2015).

### List of current members of the Genes & Health Research Team

**Aston University**

Eamonn Maher,

**Blizard Institute, Queen Mary University of London**

Shabana Chaudhary,

Joseph Gafton,

Karen A Hunt,

Shapna Hussain,

Kamrul Islam,

Mohammed Bodrul Mazid,

Elizabeth Owor,

Jessry Russell,

Nishat Safa,

John Solly,

Marie Spreckley,

David A Van Heel,

Jan Whalley,

Ishevanhu Zengeya,

Emily Mantle,

**Bradford Teaching Hospitals NHS Foundation Trust**

Shaheen Akhtar,

Samina Ashraf,

Dan Mason,

John Wright,

**Garvan Institute**

Daniel MacArthur,

**King’s College London**

Michael Simpson,

Richard C Trembath,

Gerome Breen,

Raymond Chung,

Sang Hyuck Lee,

**Manchester University Hospitals**

Omar Asgar,

Joanne Harvey,

Karen Tricker,

Caroline Winckley,

Hanifa Khatun,

Amna Asif,

**Precision Healthcare University Research Institute, Queen Mary University of London**

Claudia Langenberg,

**Social Action for Health (charity)**

Grainne Colligan,

Ceri Durham,

**University of Manchester**

Bill Newman,

**Waltham Forest Council**

Ahsan Khan,

**Wellcome Sanger Institute**

Hilary Martin,

Teng Heng,

Matt Hurles,

Vivek Iyer,

Georgios Kalantzis,

Vladimir Ovchinnikov,

Iaroslav Popov,

Klaudia Walter,

**William Harvey Research Institute, Queen Mary University of London**

Panos Deloukas,

David Collier,

**Wolfson Institute of Population Health, Queen Mary University of London**

Ana Angel,

Saeed Bidi,

Fabiola Eto,

Sarah Finer,

Chris Griffiths,

Sam Hodgson,

Benjamin M Jacobs,

Rohini Mathur,

Caroline Morton,

Asma Qureshi,

Stuart Rison,

Annum Salman,

Miriam Samuel,

Moneeza K Siddiqui,

Daniel Stow,

Sabina Yasmin,

Julia Zöllner,

Sheik Dowlut,
